# PFAS Profiles in Private Wells Reveal Septic System Sources of Contamination in a Rural Maine Community

**DOI:** 10.64898/2026.10.06.26364605

**Authors:** Richard F. Hilliard, Emma McDonald, Léo Charbonneau, Caitlin Oliver-Olsen, Alejandro M. Fernández Snell, Alexis C. Garretson, Jane E. Disney

## Abstract

Per- and polyfluoroalkyl substances (PFAS) are present throughout the environment due to decades of use in many commercial products and industrial processes. These molecules contain a hydrophobic fluorocarbon tail and a functional headgroup (typically a carboxylic or sulfonic acid) that allows hydrophilic interactions with the surrounding environment. These properties make them inherently resistant to degradation, difficult to track through the environment, readily bioaccumulative, and likely to impact human health. Through collaboration with town officials, multiple meetings with local sustainability committee members and school officials, and direct contact with individual homeowners, we conducted community-driven sampling of PFAS in well water from 105 houses in Tremont, ME, a rural coastal town that relies on private wells for drinking water. The rate of PFAS detection for one or more analytes across all wells (N=105) was 69%. Of all wells tested, 16 (15%) exceeded the current public drinking water standard in Maine (20 ng/L for 6 PFAS), and an additional 17 (totaling 31% of all wells tested) exceeded Maine’s newly adopted public drinking water standards for the state (based on EPA 2024 standards). Because private wells are unregulated, homeowners may use these results as a guide for taking action to maintain safe drinking water. Both hierarchical cluster analyses and pairwise Spearman’s rank correlation of PFAS test results revealed similar profiles in households near and far from one another. Along with the absence of obvious, large-scale potential point sources of PFAS contamination, these findings support septic systems as the primary source of PFAS contamination in drinking water across this town. The community-driven approach to the sampling effort has enabled information sharing on the importance of water filtration options, discussion of long-term pollution abatement strategies for individual households and the town, and focused efforts for future well water testing and PFAS mitigation.

**Table of Contents Graphic / Abstract Graphic:** 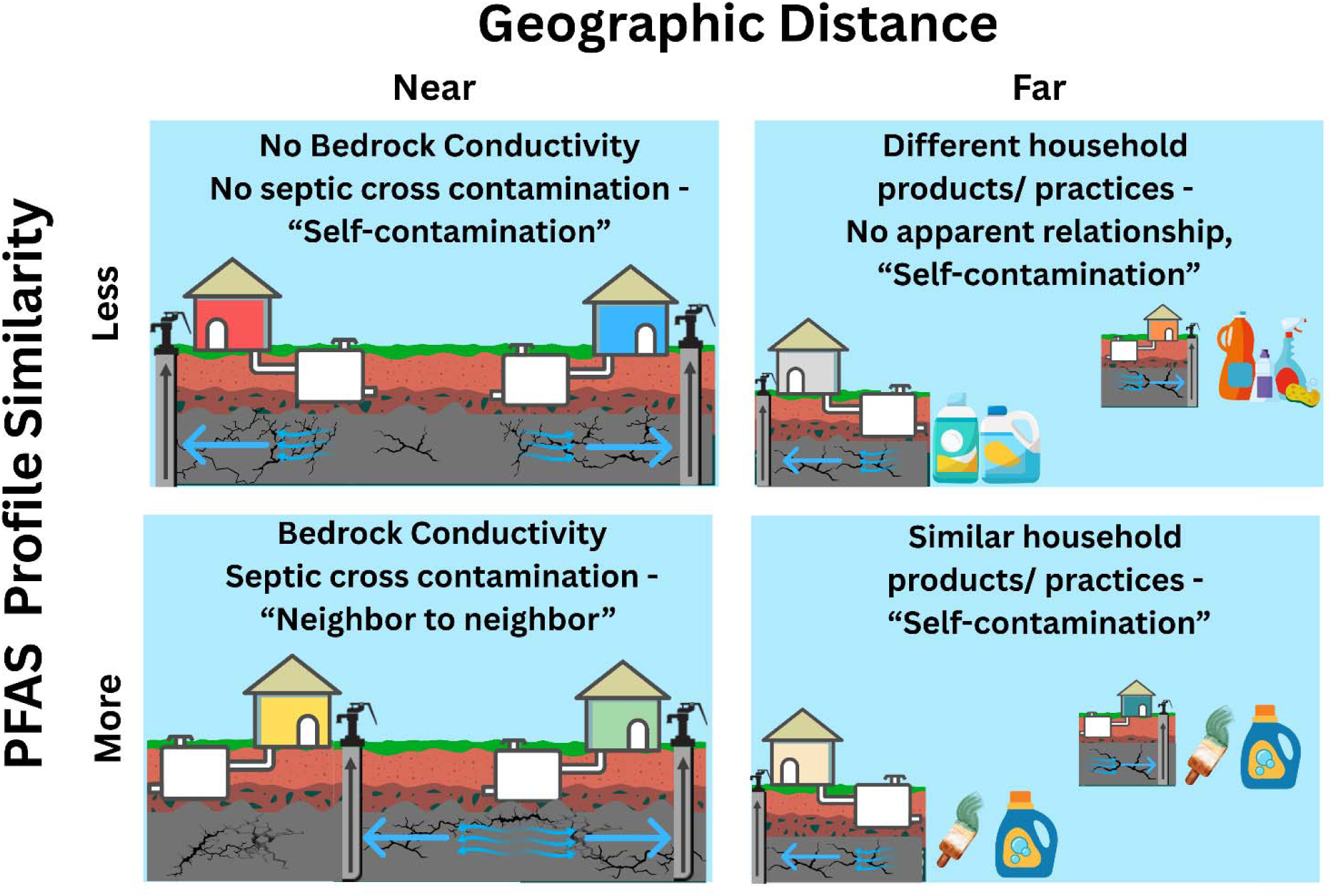

## Introduction

Per- and polyfluoroalkyl substances (PFAS) are a class of synthetic organofluorine compounds produced since the mid-twentieth century that resist degradation, bioaccumulate, and are associated with human health issues; they are now widespread in people, wildlife, and the natural environment. These chemicals are defined by the Organization for Economic Co-operation and Development (OECD) as substances that contain at least one fully fluorinated methyl [-CF_3_] or methylene [-CF_2_-] carbon atom (Schymanski *et al.,* 2023). These carbon-fluorine bonds are very strong, approximately 485 kJ/mol (Puts, Crouse and Ameduri, 2019), making these compounds incredibly resistant to degradation. These chemicals are incorporated into products and processes to meet as many as 325 different applications across 18 distinct use categories; including, within the context of a single residence: cookware; food packaging; cleaning; waterproofing; and repelling of water, oils, and greases (Figuière *et al.,* 2025). Because they are used in a wide range of consumer products and industrial processes without proper safeguards for disposal or pollution prevention, these compounds are currently detected in numerous environmental matrices, including air, soil, surface water, and groundwater.

Mechanistic understanding of the health impacts of PFAS is still developing, but it is broadly accepted that PFAS are linked to kidney and testicular cancer, metabolic disruption including increased liver enzymes and cholesterol, immune dysfunction, lower birth weights, shorter breastfeeding times, and endocrine system disruptions (DeWitt *et al.,* 2026). An estimated 61% of human PFAS exposure is attributable to drinking water contamination (Wood *et al.,* 2025; Boettger *et al.,* 2025). To address the public health implications of PFAS exposure via drinking water, we need a better understanding of how these chemicals enter drinking water sources.

Increasing rates of PFAS testing and advances in analytical techniques have revealed widespread contamination of drinking water supplies. PFAS are regularly detected in groundwater in the eastern United States, with 20% of wells having at least one analyte (McMahon *et al.,* 2022). Some are as high as 5,200 ng/L (Hepburn *et al.,* 2019). Across the province of Quebec, including public and private sources from 376 municipalities, 99.3% of samples had at least one detectable analyte and higher summations of PFAS compounds in surface water than groundwater, but 6 of the 10 most contaminated samples were from groundwater, with two outlier sites (with unusually high concentrations) related to groundwater (Munoz *et al.,* 2023). In a Pennsylvania study, 65% of 167 private wells had detectable PFAS, with two legacy compounds, perfluorooctane sulfonic acid (PFOS, 52%) and perfluorooctanoic acid (PFOA, 47%), detected most frequently, and 18% of wells exceeded at least one health-based standard (Kosiarski *et al.,* 2025). A nationwide review estimated that at least one PFAS compound would be detected in 45% of US drinking water samples (Smalling *et al.,* 2023), again with PFOS and PFOA most likely to be detected. In a Wisconsin study, 71% of samples from shallow private wells had at least one detectable PFAS (Silver *et al.,* 2023).

Contamination of groundwater by PFAS has been attributed to numerous sources, including production and manufacturing, especially chemical industries (Davis *et al.,* 2007; Giglioli, Colombo and Azzellino, 2023; Jensen *et al.,* 2024; Teymoorian *et al.,* 2025; Whitehead *et al.,* 2025); sites like airports, military bases, and fire stations where aqueous film-forming foam (AFFF) has been used to suppress fires (Houtz *et al.,* 2013; Reinikainen *et al.,* 2022; Nason *et al.,* 2024, 2024; Alexander *et al.,* 2025; Ashworth *et al.,* 2026), and landfills (Hepburn *et al.,* 2019). Numerous studies also document areas of city- and municipal-scale that have multiple sources of PFAS contamination (Giglioli, Colombo and Azzellino, 2023; Kim *et al.,* 2025), with distinct chemical signatures observed across different land use types within one city (Sun *et al.,* 2025). Even small municipalities may have contaminated groundwater attributable to multiple sources, for example, both a firefighting station and municipal and domestic wastewater treatment (Weber *et al.,* 2017); or both a landfill and a military base (Sadia *et al.,* 2023).

Wastewater is increasingly recognized as a source of PFAS in groundwater. Biosolids, the solid byproducts of wastewater, whether used for agriculture or fill, are a known source of PFAS in the subsurface (Johnson, 2022). Even small- and medium-scale wastewater systems have been documented as sources of PFAS (Lin *et al.,* 2024) which may impact nearby wells (Moran Sosa *et al.,* 2024; Penrose *et al.,* 2025). Groundwater near reclaimed water recharge facilities can be impacted (Cáñez *et al.,* 2021) and rural schools can contaminate residential drinking water wells in their proximity (Moran Sosa *et al.,* 2024). Importantly, PFAS contamination of septic systems has been associated with co-contaminants such as nitrate and pharmaceuticals (Schaider, Ackerman and Rudel, 2016; Joseph *et al.,* 2025). In shallow wells in Wisconsin, PFAS contamination was associated with signs of septic impacts via developed land and human waste indicators (artificial sweeteners and pharmaceuticals), and in some cases, agricultural waste (Silver *et al.,* 2023). However, systematic understanding of septic systems as sources of PFAS remains limited (Gyimah *et al.,* 2024) and is only now emerging (Harfmann *et al.,* 2026).

Across the United States, about 1 in every 5 households use individual or small community septic systems to treat wastewater, and in more rural regions like New England, this rate approaches 3 in every 10 (Yu *et al.,* 2024). As PFAS contamination in groundwater may be attributed to both wastewater treatment (Cáñez *et al.,* 2021; Penrose *et al.,* 2025) and land-applied sludge and septage (Moavenzadeh Ghaznavi *et al.,* 2023), it is critical to consider septic systems as sources of PFAS in groundwater (Schaider, Ackerman and Rudel, 2016).

In Maine, broad concern about PFAS traces to the detection of PFAS in dairy milk, and extended quickly to sites where biosolids, sewage sludge, and septage had been land-applied, often on farms, but also on areas where fill was required, such as sports fields and construction sites (Moavenzadeh Ghaznavi *et al.,* 2023). As in other states (Peter and Lee, 2025), biosolids have been widely applied to agricultural fields in Maine, contributing to PFAS detection in drinking water from private wells and correlating with levels in blood serum of affected homeowners (Criswell *et al.,* 2025).

An estimated 17% of the United States population uses private wells for drinking water (Murray *et al.,* 2021), but in Maine, 55% of households rely on private wells (Maine Tracking Network, 2023). PFAS are regulated in public water supplies in Maine; this includes wells having “at least 15 service connections” or serving “at least 25 individuals daily at least 60 days out of the year” (*Maine Revised Statute. Title 22, §2601(8)*). These are regulated to a standard—referred to here as ME-6—of no more than 20 ng/L total for a summation of six individual PFAS compounds: PFOS and PFOA, along with perfluorohexane sulfonic acid (PFHxS), perfluoroheptanoic acid (PFHpA), perfluorononanoic acid (PFNA), and perfluorodecanoic acid (PFDA) (S.P. 64 - L.D. 129, “*Resolve, To Protect Consumers of Public Drinking Water by Establishing Maximum Contaminant Levels for Certain Substances and Contaminants*”). Beginning in April 2027, public water systems in Maine will begin monitoring under the federal rule—referred to here as EPA-2024—and by April 2029 will be in full compliance with this more stringent standard of no more than 4 ng/L of either PFOA or PFOS, no more than 10 ng/L of PFHxS, PFNA, or hexafluoropropylene oxide dimer acid (HFPO-DA), and a Hazard Index no larger than 1(*Maine Revised Statute. Title 22, §2650-A(2)*).

In 2024, we responded to community concerns about PFAS contamination of drinking water at a local high school by initiating a well testing project in the neighborhood surrounding the school (Moran Sosa *et al.,* 2024). This effort also prompted the nearby Tremont community to take an interest in well water testing for PFAS.

The town of Tremont was familiar with PFAS as a drinking water issue. In 2017, PFAS and other persistent organic pollutants were detected in two on-site monitoring wells around a municipal landfill that had closed in 1996. Consequently, PFAS were discovered in nine private drinking water wells on nearby properties. The Maine Department of Environmental Protection (DEP) tested and remediated these private wells with activated carbon filters, which the town now maintains. The Maine Department of Environmental Protection (DEP) considers the groundwater contamination to be confined to areas directly adjacent to this closed landfill site.

When homeowners in Tremont heard about the well testing near the high school, they reached out to us for support in testing their own wells. The town select board supported this effort by funding 50 household tests. A Maine-based commercial PFAS analysis laboratory provided a discounted price on PFAS tests. With additional grant funding, we collected well water samples from 105 households; 69% had detectable PFAS, and 15% exceeded the Maine standard of 20 ng/L for 6 PFAS. We looked for patterns and relationships between and among PFAS profiles to infer potential contamination sources so people could take action to protect their health. To accomplish this, we sorted each household with detectable PFAS into a hierarchical cluster and found four groups dominated by a particular PFAS type. Households within clusters were not necessarily close geographically, suggesting something similar happened at each location to contaminate the wells. Based on these profiles, and the lack of obvious nearby sources of PFAS, we theorize that most of these households may have “self-contaminated” their wells through past or present use of household products containing PFAS, which entered the septic system and leach fields and percolated into bedrock aquifers (found throughout Maine) used for drinking water. Neighboring wells with similar chemical profiles may show cross-contamination instead of, or in addition to, “self-contamination”. These findings warrant further investigation and demonstrate a need for PFAS testing of drinking water in rural communities even where there is no obvious potential source of PFAS contamination.

## Methods

### Site Description

This study was conducted in the town of Tremont located in Hancock County, Maine. Tremont, a coastal rural town of approximately 1500 residents, is on Mount Desert Island in the Gulf of Maine, near the midpoint of Maine’s fractal coastline. The borders of this town are formed by the sea to the east and south and preserved forest and marsh to the north and west. Like many rural communities, this town has no public water system; as such, each residence relies on a private well.

### Study design

This study, conducted between 2023 and 2026, used a participatory framework in which residents, homeowners, and property owners expressed interest in a study, attended community meetings to discuss next steps, and signed up to have their well water tested for PFAS. We distributed flyers with a QR code and a link to a Google Form to collect names, addresses, email contacts, dates and times of availability, mailing address, phone number, comments, and interest in a PFAS test. We partnered with the Tremont Sustainability Committee at the recommendation of the Tremont Town Manager; they helped share information, distribute flyers, and host multiple public meetings at the local library. We contacted homeowners to schedule visits to their homes, where we collected water samples. Homeowners also provided, when available, information about well type, well depth, filtration status, and other associated details.

The relationships between the researchers and the town Selectboard, Sustainability Committee, school administrators, and individual residents continued to evolve through numerous community meetings, presentations, emails, and house visits. Community members have been regularly updated on preliminary findings, advised on filtration options, informed about what is known and what remains unknown regarding PFAS and their health, and updated on changing state and federal standards regulating PFAS in drinking water.

### Sample collection

Drinking water samples were collected using kits provided by PFAS Laboratories (formerly Maine Laboratories) in Norridgewock, Maine, containing PFAS-free supplies, like Ziplock® bags and high-density polyethylene (HDPE) bottles, (Wanzek *et al.,* 2024). Kits were comprised of a one-gallon Ziplock® bag containing three smaller Ziplock® bags, one with a chain of custody form and clean single-use gloves, another with two 250 mL sample bottles for primary and duplicate samples, and a third with an empty 250 mL bottle labeled "Field Reagent Blank " and another 250 mL bottle labeled "Lab Reagent Water” with PFAS-free water to pour into the field blank bottle during sampling. All bottles used were high-density polyethylene (HDPE). Before sampling, we opened faucets to about one-third capacity and ran them at a moderate flow rate for 5 minutes. We filled sample bottles to the indicated line (250 mL) and inverted them 5 times to mix. When possible, we bypassed household filters at the time of sample collection. In some cases, sediment filters, which are not effective for PFAS removal, were not bypassed. The Supplemental Information includes filter use and type. Samples, duplicates, and field blanks were stored at 4 °C, packed in coolers with ice, picked up by a courier service, and delivered within 24 hours to PFAS Laboratories, where they were analyzed within two weeks of collection.

### Sample Analysis for PFAS

PFAS analyses of 28 analytes were performed by PFAS Laboratories in Norridgewock, ME, using method 537 MOD (Shoemaker, Grimmett and Boutin, 2008). Briefly, a 250 mL water sample is fortified with isotopic surrogates and passed through a solid-phase extraction cartridge containing polystyrene-divinylbenzene. The analytes and surrogates are eluted with a small amount of methanol. This extract is dried with nitrogen in a heated water bath, then adjusted to 1 mL after adding internal standards. A 10 µL aliquot is then injected into a liquid chromatography (LC) column (C_18_ spec). This separates the analytes and sends them to a tandem mass spectrometer (MS/MS), which uses two stages of ionization and mass-to-charge ratio measurements to quantify the analytes.

#### Data Filtering

For data analysis, the dataset was trimmed to include only analytes that were detected in 3 or more samples, those analytes are: perfluorobutanoic acid (PFBA), perfluoropentanoic acid (PFPeA), perfluorobutanesulfonic acid (PFBS), perflourorhexanoic acid (PFHxA), perfluorohexanesulfonic acid (PFHxS), perfluoroheptanoic acid (PFHpA), perfluorooctanoic acid (PFOA), perflurooctanesulfonate (PFOS), perfluorononanoic acid (PFNA), and perfluorodecanoic acid (PFDA). These analytes account for 98.7% of the total mass measured. The following analytes which were never present above the detection limit in any sample were trimmed from the dataset: 4:2 fluorotelomer sulfonate (4:2 FTS), 2,3,3,3-Tetrafluoro-2-(heptafluoropropoxy)propionic acid (HFPO-DA also known as GenX), 4,8-Dioxa-3H-perfluorononanoic acid (ADONA), perfluorohetpanesulfonic acid (PFHpS), 2-((6-Chloro-1,1,2,2,3,3,4,4,5,5,6,6-dodecafluorohexyl)oxy)-1,1,2,2-tetrafluoroethanesulfonic acid (9Cl-PF3ONS), 1H,1H,2H,2H-Perfluorodecanesulfonic acid (8:2FTS), perfluoroundecanoic acid (PFUnA), 11-Chloroperfluoro-3-oxaundecanesulfonic acid (11Cl-PF3OUdS), PFDoA (perfluorododecanoic acid), PFNS (perfluorononanesulfonic acid), NMeFOSAA (2-(N-Methylperfluorooctanesulfonamido)acetic acid), PFOSA (Perfluorooctanesulfonamide), PFDS (perfluorodecanesulfonic acid), PFTrDA (perfluorotridecanoic acid), and PFTeA (perfluorotetradecanoic acid). Finally, one analyte was detected once, NEtFOSAA (N-Ethyl-N-[(heptadecafluorooctyl)sulphonyl]glycine) at 1.55 ng/L; two analytes, PFPeS (Perfluoropentanesulfonic acid; at 0.30 and 0.39 ng/L) and 6:2 FTS (6:2 Fluorotelomer sulfonic acid; at 0.98 and 3.24 ng/L) were detected twice; these were also trimmed from the dataset, but are available in the SI.

#### Data Management, Storage and Availability

PFAS Laboratories sent water test reports to each homeowner. We also received copies of the reports and an Excel spreadsheet with compiled PFAS data. Well water information and associated PFAS data are stored on the anecdata.org website developed by the Community Environmental Health Lab at MDI Biological Laboratory (Bailey *et al.,* 2021; Garretson *et al.,* 2023). The dataset is available in Supplementary Information, with personal identifying information removed.

#### Data Analyses

Analyses were performed on an Apple computer running macOS Sequoia 15.6 with a 2 GHz Quad-Core Intel Core i5 processor. **Summary Statistics.** Summary statistics (mean, percentiles, population standard deviation, range, total, and fractional rate of detection) for all sites were calculated using RStudio Version 2025.05.1+513 and R version 4.5.3. "Reassured Reassurer").

#### Hierarchical Clustering

Hierarchical clustering was performed including only samples with any detectable PFAS, using a ‘Euclidean’ distance matrix and ‘average’ clustering method with the ‘hclust’ package, and output with ‘pheatmap’ (version 1.0.13). We also tested additional clustering methods: ward.D, ward.D2, single, complete, mcquitty, median, and centroid. Prior to cluster analysis, PFAS data were centered by subtracting the mean analyte concentration from each given measurement:

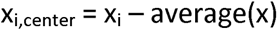

and were scaled, normalizing each measurement by the root mean squared value for that analyte:

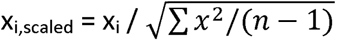

#### Spearman’s rank correlation

We calculated pairwise correlations of samples (only those with any detected PFAS) as Spearman’s rho coefficients based on the rank order of individual analytes by concentration. Non-detects of individual analytes are treated as concentrations of 0.0 ng/L.

#### Principal Component Analysis

A principal component analysis (PCA) was performed using centered and scale data (as described above) using the prcomp function in the “stats” package (version 4.5.3) for R.

#### Geographic Designations and Mapping

This municipality has several historical maritime villages, which we approximate as 4 zones (see **Figure 1**). The boundaries between these villages are not universally agreed upon, so we used simplified designations labeled A, B, C, and D. We assigned samples to Zones via a point-in-polygon merge using a shapefile of these zones and the latitude and longitude of each site. Each site was reverse-geocoded to latitude and longitude from the residential street address using OpenStreetMap’s Nominatim service and was manually verified when data were entered into anecdata.org.

**Figure 1.**
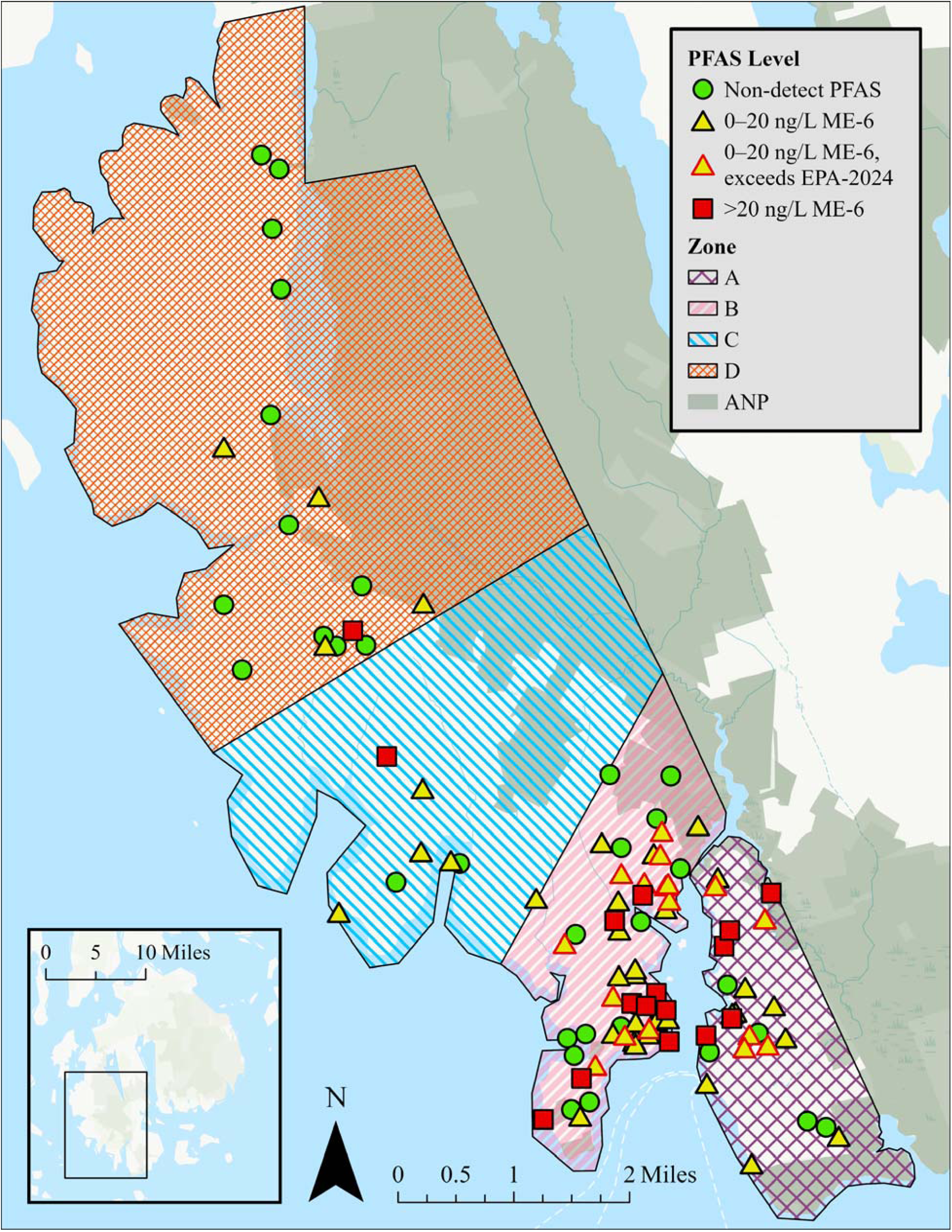
Map of private wells tested for PFAS. All locations have been obfuscated as described in the Methods section. Green dots = no PFAS detected. Yellow triangles = any detected PFAS and below 20 ng/L for the ME-6. Red = above 20 ng/L for the ME-6. Yellow triangles with a red border exceed the EPA-2024 standard, which will be enforced for public water systems in Maine by 2029. ‘ANP’ designates Acadia National Park.

#### Obfuscation

On all maps, we obfuscated sample sites by offsetting coordinates. We derived offset locations by adding random values between -0.004 and 0.004 to the decimal latitude and longitude. The average offset across all sites was 282.5 meters, with a minimum and maximum offset of 59.4 and 496.7 meters, respectively. On any map where an individual site is located at or near a residence, that is not the true location where the water sample was collected.

**Geographic clustering** was performed using pairwise distances between each point. These distances were calculated using a Haversine distance from the geosphere package (1.5-20), based on latitude/longitude and the default radius of r = 6,378,137 meters. The pairwise distances (in meters) were matched by the intersection of the site (row name) with the columns labeled with the other site names (e.g., ‘A-2’, ‘D-13’). Within these sectors, we randomly numbered sites. We created maps using ArcGIS Pro 3.6.2 on a 2.11 GHz Intel® Core™ i7 processor running Windows 11 Pro.

#### Population Density Estimation

We used the number of tax parcels as a proxy for population density. We obtained a map of tax parcels for the town from the State of Maine’s “Maine Digital Parcel Viewer Web Map” and, using ArcGIS Pro, counted the number of parcels per zone, then normalized it by dividing by the area of that Zone.

## Results and Discussion

The rate of PFAS detection across all private wells (N=105) for one or more analytes in this municipal-scale study was 69%. This is a higher percentage than the broad Eastern US, which has been estimated at 20% (McMahon *et al.,* 2022), higher than the estimated 45% of US drinking water samples, including both public and private supplies (Smalling *et al.,* 2023), but similar to the statewide survey of shallow drinking water wells in Wisconsin, USA (Silver *et al.,* 2023) in which 71% of wells had at least one detectable PFAS analyte.

**Figure 1** shows the approximate location of each well. Green dots indicate households with no detectable PFAS (n=33); yellow triangles indicate those with detectable PFAS levels below the ME-6 standard (n=56); and red squares indicate those that exceed ME-6 (n=16). Yellow triangles with red borders (n=17) indicate the subset of samples below ME-6 that exceed EPA-2024, which Maine adopted in 2025 and will enforce by 2029, while yellow triangles with black borders remain below this new standard. **Table 1** lists the summary statistics for this study. PFOA (62%) and PFBA (57%) were detected most often. While PFOA is regulated in public drinking water, PFBA, which accounted for 8.5% of detected mass, is not included in either the ME-6 or EPA-2024 standards. Five additional analytes (PFPeA, PFBS, PFHxA, PFHpA, and PFOS) were each detected in more than 44% of the samples. Three of these analytes, PFBS, PFPeA, and PFHxA, are not included in the current state drinking water standard (ME-6). Shortly, Maine will enforce the EPA-2024 standard, which will apply restrictions for PFBS, but only as a component of the Hazard Index calculation discussed above (a water source contaminated with PFBS but not PFNA, PFHxS, or HFPO-DA (Gen-X) would have to have over 2,000 ng/L PFBS to be in violation of the Hazard Index provision of the standard (Maine Revised Statute, Title 22, §2650-A(2)).

**Table 1.**
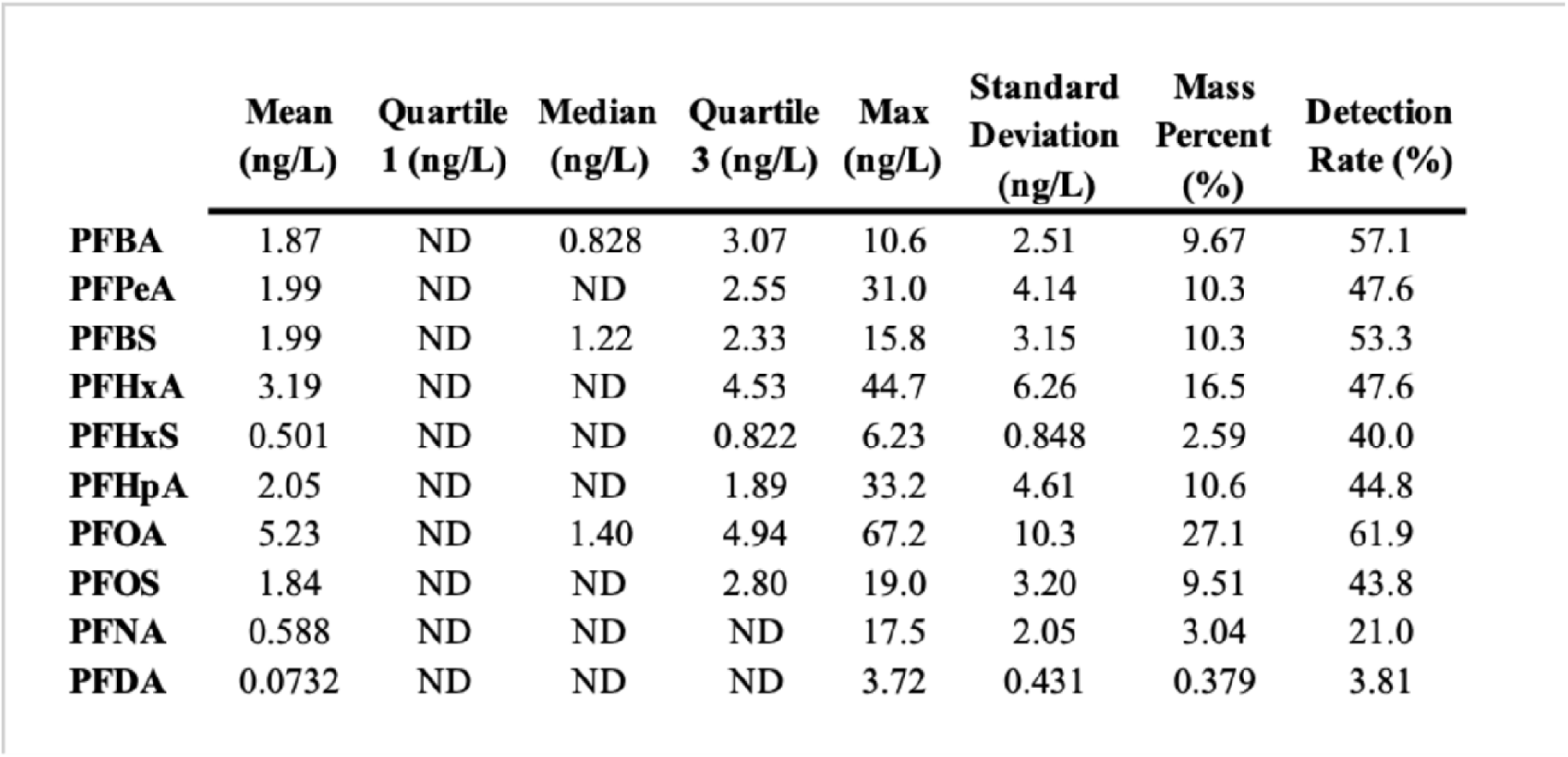
Summary statistics of PFAS measurements across the town of Tremont

|  | Mean<br>(ng/L) | Quartile<br>1 (ng/L) | Median<br>(ng/L) | Quartile<br>3 (ng/L) | Max<br>(ng/L) | Standard<br>Deviation<br>(ng/L) | Mass<br>Percent<br>(%) | Detection<br>Rate (%) |
| --- | --- | --- | --- | --- | --- | --- | --- | --- |
| **PFBA** | 1.87 | ND | 0.828 | 3.07 | 10.6 | 2.51 | 9.67 | 57.1 |
| **PFPeA** | 1.99 | ND | ND | 2.55 | 31.0 | 4.14 | 10.3 | 47.6 |
| **PFBS** | 1.99 | ND | 1.22 | 2.33 | 15.8 | 3.15 | 10.3 | 53.3 |
| **PFHxA** | 3.19 | ND | ND | 4.53 | 44.7 | 6.26 | 16.5 | 47.6 |
| **PFHxS** | 0.501 | ND | ND | 0.822 | 6.23 | 0.848 | 2.59 | 40.0 |
| **PFHpA** | 2.05 | ND | ND | 1.89 | 33.2 | 4.61 | 10.6 | 44.8 |
| **PFOA** | 5.23 | ND | 1.40 | 4.94 | 67.2 | 10.3 | 27.1 | 61.9 |
| **PFOS** | 1.84 | ND | ND | 2.80 | 19.0 | 3.20 | 9.51 | 43.8 |
| **PFNA** | 0.588 | ND | ND | ND | 17.5 | 2.05 | 3.04 | 21.0 |
| **PFDA** | 0.0732 | ND | ND | ND | 3.72 | 0.431 | 0.379 | 3.81 |

The highest total PFAS level detected in this study was 159 ng/L, with 101 ng/L (ME-6) in an approximately 300 ft deep well in Zone B. This maximum total is far below levels near biosolids-impacted areas, where concentrations as high as 24,000 ng/L have been reported (Criswell *et al.,* 2025) but is remarkable because this well is not located near any known industrial or commercial sites. The average total PFAS across all wells was 19.4 ng/L when including non-detects as zeros, with a median total PFAS of 7.18 ng/L (excluding non-detects, the average total PFAS was 28.2 ng/L).

PFOA was not only the most frequently detected compound, but it also accounted for approximately 27% of the total PFAS mass recovered from all samples. PFHxA accounts for 14.9%, and PFBA, PFPeA, PFBS, PFHpA, and PFOS each account for about 10% of the overall mass. Three analytes: PFHxA, PFBA, and PFPeA do not fall under the current ME-6 framework or the future EPA-2024 regulations, but are all detected in more than 48% of samples and account for more than 36% of the mass of PFAS measured.

Two analytes, PFHpA and PFDA, regulated under the ME-6 standard, will no longer be included in the EPA-2024 regulation. For PFHpA, toxicity notwithstanding, regulation may be obviated because it is frequently associated with other PFAS, namely PFOA. In this study, PFHpA was never detected without PFOA, and therefore may be of secondary concern for regulation. Similarly, PFDA was never detected in this study without numerous other PFAS present and accounts for less than 0.4% of the overall mass detected. PFDA detection from all samples in this study totaled 7.7 ng/L. This low level of detection suggests that PFDA may not be as critical a regulatory concern. While 69% of wells had some detectable PFAS, the data largely consist of non-detect analytes. Of the ten analytes appearing in three or more well water samples, only 3 of these recorded a median value above non-detect: PFBA, PFBS, and PFOA. For all analytes, the 25^th^ percentile value is non-detect. Although even rural areas are at risk of PFAS-impacted groundwater, improvements in analytical methods and revisions toward more stringent drinking water standards support continued monitoring in areas already examined for PFAS contamination.

### Chemical Profiles

The dendrogram in **Figure 2** shows the hierarchical cluster analysis of scaled profiles for each site, revealing the central importance of legacy pollutants (PFOA and PFOS) in clustered associations, as well as the predominance of PFHxA and PFBS. These clusters of related household drinking water samples are driven primarily by these four PFAS, indicated by framed groupings of red cells in the dendrogram. The branch groupings are driven by two carboxylic acids, PFOA and PFHxA, on one branch, and two sulfonic acids, PFOS and PFBS, on another branch. Furthermore, repeated cluster analyses using different algorithms (ward.D, ward.D2, single, complete, mcquitty, median, centroid) also group PFOA and PFHxA together, and PFOS and PFBS together. This aligns with previous groundwater observations, where similar PFAS (e.g., carboxylates like PFHpA, PFHxA, and PFBA) are more likely to be detected alongside the legacy carboxylate PFOA than PFOS (Silver *et al.,* 2023).

**Figure 2.**
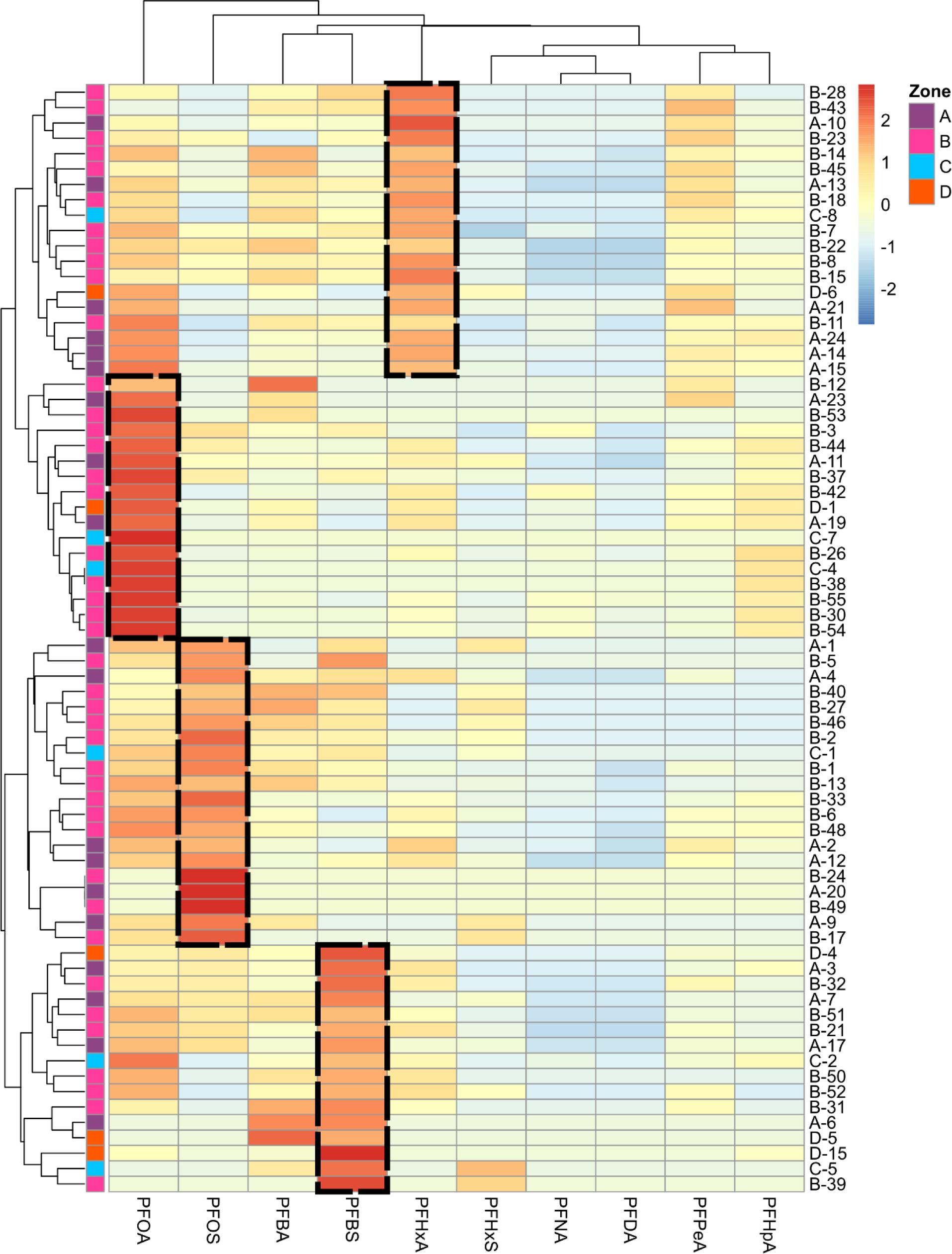
Cluster analysis of PFAS profiles measured in drinking water (n=72) wells from a single rural Maine town. Household drinking water samples are labeled along the right-hand side. A dendrogram on the left shows each branch with two sub-branches, indicating related PFAS distributions in well water samples. Color intensity (red = positive, blue = negative) indicates the influence of the individual PFAS analyte (along the bottom) in clustering a particular site within the dendrogram(tree). Clusters are identified by black outlines. One black box indicates the ‘PFHxA’ cluster, which is examined in more detail in Figure 3.

**Figure S1** shows the strong associations between carboxyl and sulfonyl groups, as well as the likelihood of finding a particular PFAS alongside another PFAS. Dimension 1 of the PCA (x-axis) shows that no PFAS are independent of one another; all trend in the same direction (left). This is similar to other studies, which demonstrate that the most likely association with individual PFAS analytes in groundwater is not land use, particularly, but other PFAS analytes (Silver *et al.,* 2023). However, within this highly determined system of associations, the secondary (y) axis in **Figure S1** again demonstrates how PFAS analytes group with one another: longer-chain PFCAs (carboxylic acids) often co-occur, shorter-chain PFCAs are mostly neutral, and PFSAs (sulfonic acids) group together at the opposite pole.

As with characterizations of other impacted groundwater supplies (Smalling *et al.,* 2023), contamination of wells in this study (on a mass basis) is dominated by legacy compounds like PFOA and PFOS, and secondarily by shorter versions of these carboxylic and sulfonic acids, with more carboxylates than sulfonates detected. Higher fractions of carboxylates over sulfonates have been associated with contaminated groundwater from a city with large-scale chemical industries (Teymoorian *et al.,* 2025). However, this town has no chemical industry. Historic land use or events may have contributed to contamination of wells in Tremont. We searched historical archives from the Tremont Historical Society, Mount Desert Historical Society, and Maine State Library for historic and contemporary businesses that could have contributed PFAS to groundwater or soil, and found only a few associations with the tested households. For example, at a property in Zone C with a contaminated well (80 ng/L total PFAS), previous residents had a machine shop located near the wellhead. At another site in Zone A with a contaminated well containing 57 ng/L total PFAS, a nearby closed landfill may have contaminated their well. However, as mentioned previously, the highest total PFAS concentration (159 ng/L) was documented in a Zone B well far from any known, non-septic sources.

Figure 3, S2, S3, and S4 provide a deeper analysis of each cluster identified in Figure 2. The analysis includes a geospatial assessment (which zones each house falls in), a heat map showing relationships among individual houses in the cluster, and a chemical profile depicting the chemical similarity of the sample results. In the map in **Figure 3**, the wide distribution of chemically similar profiles—in this case from the ‘PFHxA’ cluster is denoted by red hexagons. These sites are not confined to one area of town, and many other sampling sites lie between them with no detection (green dots) or belong to one of the other 3 hierarchical clusters (displayed in **S2, S3**, and **S4**). Previous work has shown that cluster analysis was successful at associating surface waters (first-order streams) with known PFAS sources, but not for groundwater (Peter and Lee, 2025).Perhaps this is because groundwater moves more slowly than surface water and has greater potential for attenuation.

**Figure 3.**
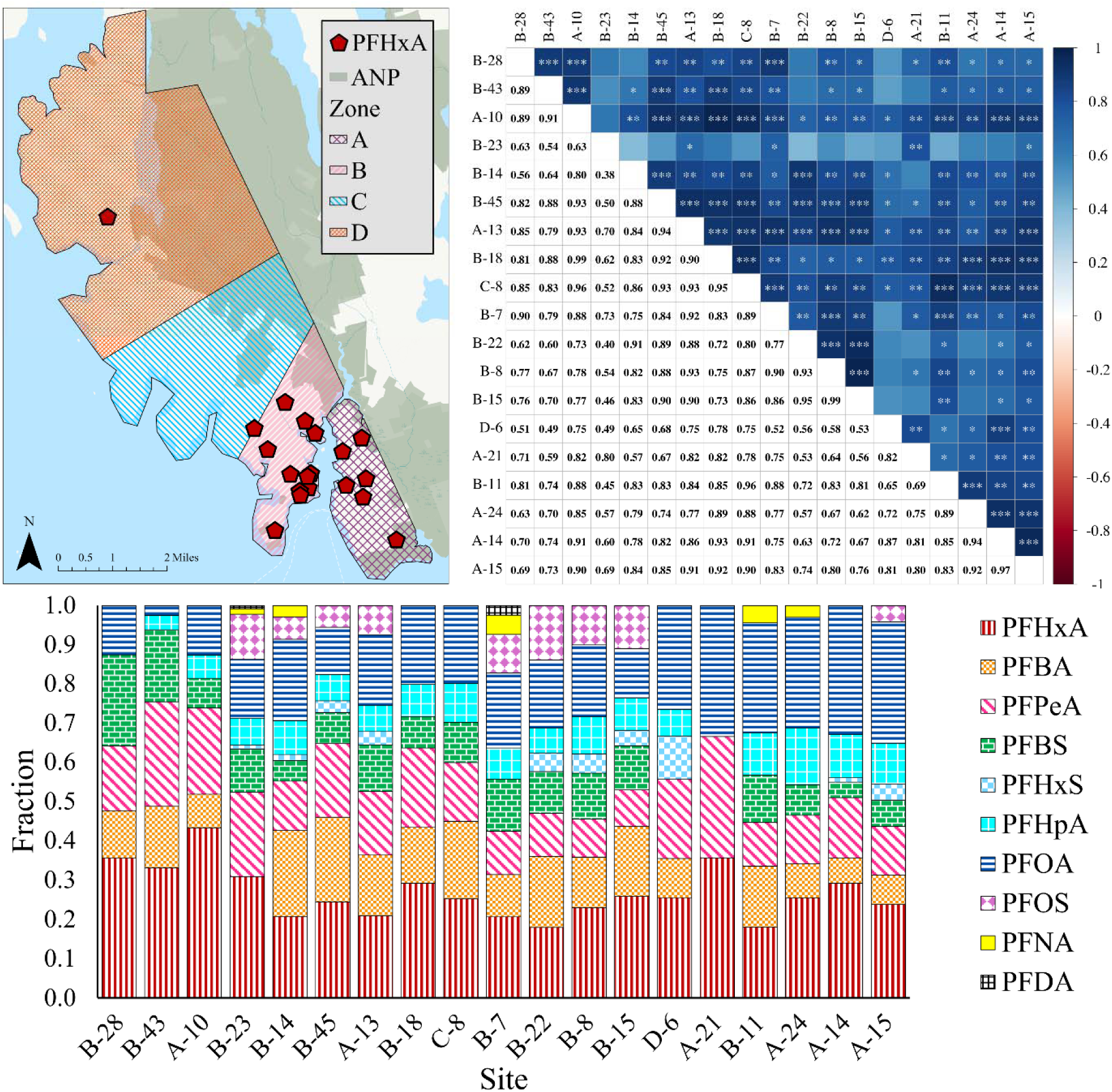
[A] Geographic distribution of wells tested for PFAS (red pentagons), [B] heat map with pairwise correlations, and [C] fractional chemical profiles of all the samples within the ‘PFHxA’ hierarchical cluster illustrated in Figure 2, by the black box surrounding 19 sites in the fifth column from the left. Each site location in 3A is offset from its true location, as described in the Methods section. In the map, ‘ANP’ designates Acadia National Park.

In **Figure S5**, a ‘tanglegram’ shows two dendrograms, each created by hierarchical clustering based on the chemical and physical distances between sites, respectively. Only three sets of samples are directly adjacent in both dendrograms. This is fewer than might be predicted, given the close proximity of many tested households (**Figure 1**). This analysis demonstrates the high likelihood of “neighbor to neighbor” contamination, and suggests that most PFAS contamination in wells is more likely due to “self-contamination” at each site, although explanations related to specific land use or bedrock geology cannot be ruled out.

Another approach to understanding PFAS contamination relationships between households across a geographic region is to do pairwise comparisons based on PFAS profiles (Moran Sosa *et al.,* 2024). Heat map analysis reveals significant correlations between households in different zones **(Figure S6)**; for example, D4 is highly correlated (P<0.001, denoted by 3 asterisks) with A-3, B32, A-7, B-21, and A-17. All of these are in the same PFHxS cluster in Figure 2, showing that PFHxS drives similarity between sites in this analysis. However, D4 is also slightly correlated with A12 (P <0 .05), which is in the PFOS cluster. This shows that the ranked order of PFAS is important for understanding relationships between contamination events across sites. In this study, correlation analysis revealed additional types and levels of relationships between households with PFAS-contaminated drinking water. Grouping sample sites by chemical similarity provides a basis for future investigations into the sources of these contaminants.

For profiles that are highly similar and from households that are physically close (such as the densely populated areas in Zones A and B), future sampling should focus on determining the connectivity between wells via possible fractures in the bedrock aquifers. For sites that are quite far apart, e.g., A-14 and D-6, yet have highly similar chemical profiles, further investigation should focus on similarities like land use; bedrock and surficial geology; as well as the use of common products; and whether residents have similar occupations or hobbies like painting, metalworking, or woodworking that might use PFAS-containing products.

**Figure 4** depicts the relationship between population density and the likelihood of PFAS detection. Zones A, B, and C have 4.8, 3.5, and 1.7 more tax parcels per area, respectively, than Zone D, the least populated zone. Zone D also has the largest fraction of wells with no PFAS detected (0.6). In contrast, in the more densely populated Zones A and B, grouped closely around the harbor in the south, more than 30% of wells in each Zone have 8 or 9 different PFAS analytes detected.

**Figure 4.**
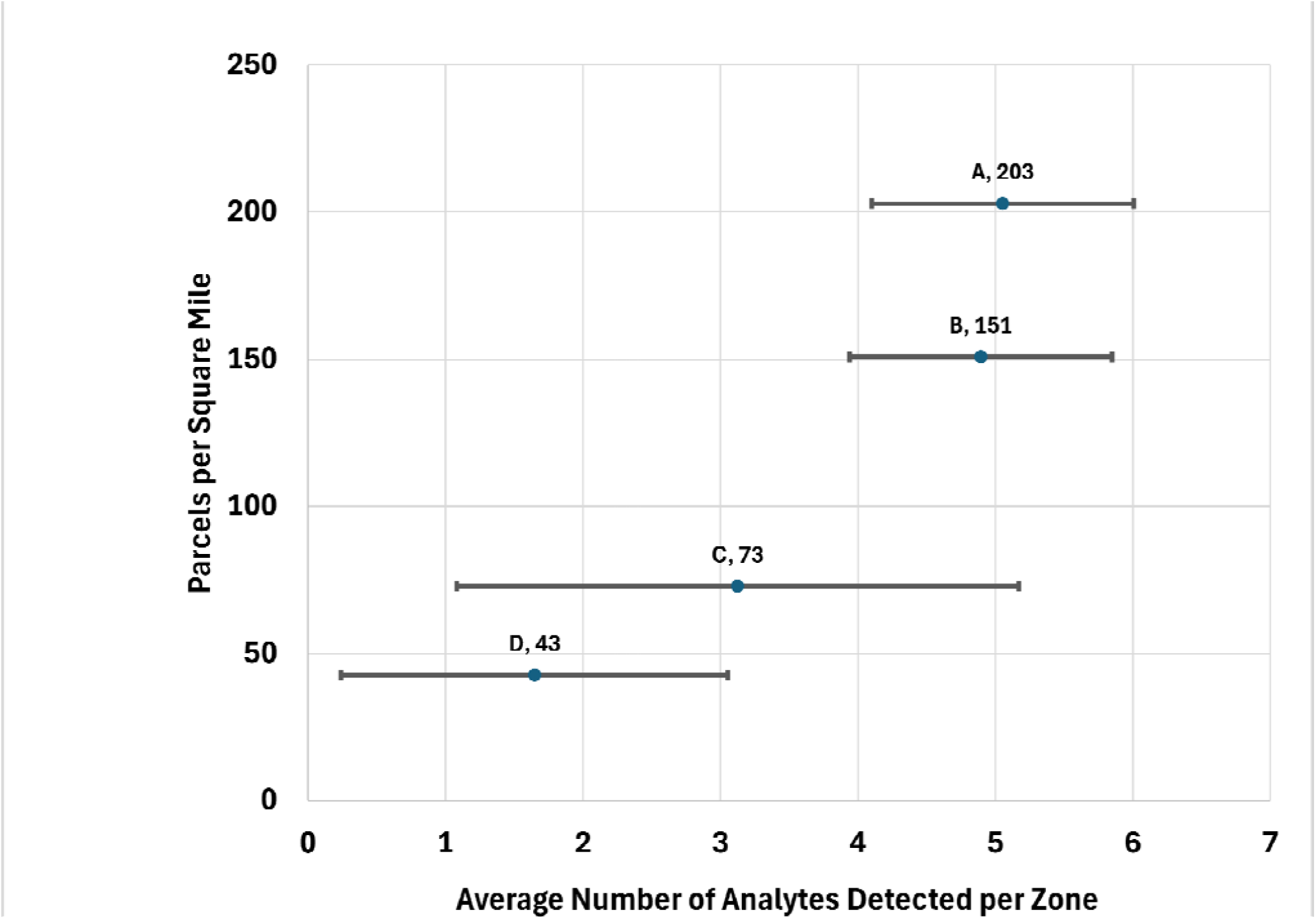
Average number of analytes detected in each geographic zone of the town, relative to the tax parcels per square mile (used as a proxy for population density). Zones are designated A, B, C, and D, based on approximate boundaries of different historic coastal settlements (see Figure 1).

The three sites in Zone D with detectable PFAS (two below ME-6, one above) are the exceptions that prove the rule. While detection rates decrease with lower population density, risks remain when residents rely on private wells. Well water testing should be a high priority in densely populated rural areas dependent on private wells for drinking water. Our municipal-scale study found a result similar to the national-scale study by Smalling et al. (2023), which found that a water sample’s location within developed land is a stronger predictor of cumulative PFAS concentration and number of detections than distance to a ‘known source’. This may signify that “neighbor to neighbor” contamination of well water via septic systems is more likely than “self-contamination” in more densely populated rural areas.

## Conclusion

PFAS contamination of groundwater will remain a serious issue for decades (Gnesda *et al.,* 2022). The fact that domestic wastewater is a documented source of PFAS (Lin *et al.,* 2024; Harfmann *et al.,* 2026), the variable character of the PFAS chemical profiles around our study area, and the clear absence of sufficient point sources, as in the cases of biosolids spreading (Holly *et al.,* 2024; Moavenzadeh Ghaznavi, 2023) raise the possibility that each individual septic system is a potential source. These systems may “self-contaminate” wells at the same residence and may also function as sources of “neighbor-to-neighbor” contamination of nearby wells. Private wells, which supply drinking water to millions of people in the US alone, and most of the land uses and residential practices that impact them, frequently fall outside both regulatory frameworks and individual awareness.

Future studies of PFAS contamination of groundwater in rural communities are critical to public health. These will be greatly aided by technological improvements in analytical methods to reach lower detection limits—as households that are presently “non-detects” may be found to have detectable PFAS—and affordable tests that allow more widespread and repeated testing. Non-target analyses (Newton *et al.,* 2025) are revealing contamination by additional PFAS which are not included in standard methods and may account for half the PFAS identified in a sample (Boettger *et al.,* 2025).

We recognize that further analysis of trace organics in domestic wells, such as pharmaceuticals or nitrates, could more conclusively indicate wastewater as the origin of PFAS in drinking water (Joseph *et al.,* 2025). Other possible sources for PFAS in groundwater in addition to septic systems include old landfills, agriculture, boat building, metal plating, or automotive repair, yet acknowledging the potential for “self-contamination” sets the stage for homeowners to take action, limit the disposal of products likely to contain PFAS down the drain, and for local ordinances and state level legislation to protect drinking water by changing land use density, septic system setbacks from wells, and limiting or prohibiting the sale of household and personal care products likely to contain PFAS (Maine Law, H.P. 1113 - L.D. 1503). Maine has already taken these steps, creating a model for the nation.

In addition to limiting the sale of products containing PFAS, states may consider conducting their own product testing, as labeling and testing of commercial and industrial products do not proceed in a central or timely manner at the federal or international level. At the local level, in addition to more widespread testing and continued monitoring over time, public health may be better informed by interviews and preliminary investigations into chemicals stored in garages and under sinks. No well, however remote from industry or other potential sources of PFAS, should be considered PFAS-free.

## Data Availability

All data produced in the present work are contained in the manuscript and supplemental tables.

## Acknowledgements

Funding for this study came from Maine IDeA Network of Biomedical Research Excellence (INBRE) and the Center for Biomedical Research Excellence (COBRE) through an IDeA from the NIGMS of the NIH under grants P20GM103423 and P20GM104318, grant P30 ES00002 from the National Institute for Environmental Health Sciences to the Harvard Chan NIEHS Center, the Fisher Charitable Foundation, the Town of Tremont, and the Morris Scientific Discovery Fund and the Shultz Fund at MDI Biological Laboratory. EKM was supported by Maine Conservation Corps as an AmeriCorps Environmental Steward on this project. This manuscript is based, in part, upon work supported by the MCC and AmeriCorps under Grant 24ESFME0010001.

## Abbreviations

AFFF: aqueous film-forming foam
DEP: Department of Environmental Protection, Maine, USA
EPA-2024: The 2024 federal standard for drinking water, denoting that no more than 4 ng/L of PFOA or PFOS, no more than 10 ng/L PFHxS, PFNA, or HFPO-DA, and a Hazard Index no larger than 1.
ME-6: The interim Maine standard for drinking water, denoting that the sum of PFOS, PFHxS, PFHpA, PFOA, PFNA and PFDA be no greater than 20 ng/L.
ng/L: nanograms per liter
PFBA: perfluorobutanoic acid
PFBS: perfluorobutane sulfonic acid
PFCAs: perfluoroalkyl carboxylic acids
PFDA: perfluorodecanoic acid
PFHxA: perfluorohexanoic acid
PFHxS: perfluorohexane sulfonic acid
PFHpA: perfluoroheptanoic acid
PFOA: perfluorooctanoic acid
PFOS: perfluorooctane sulfonic acid
PFPeA: perfluoropentanoic acid
PFNA: perfluorononanoic acid
PFSAs: perfluorosulfonic acids

## Supplemental Figures

**Figure S1.**
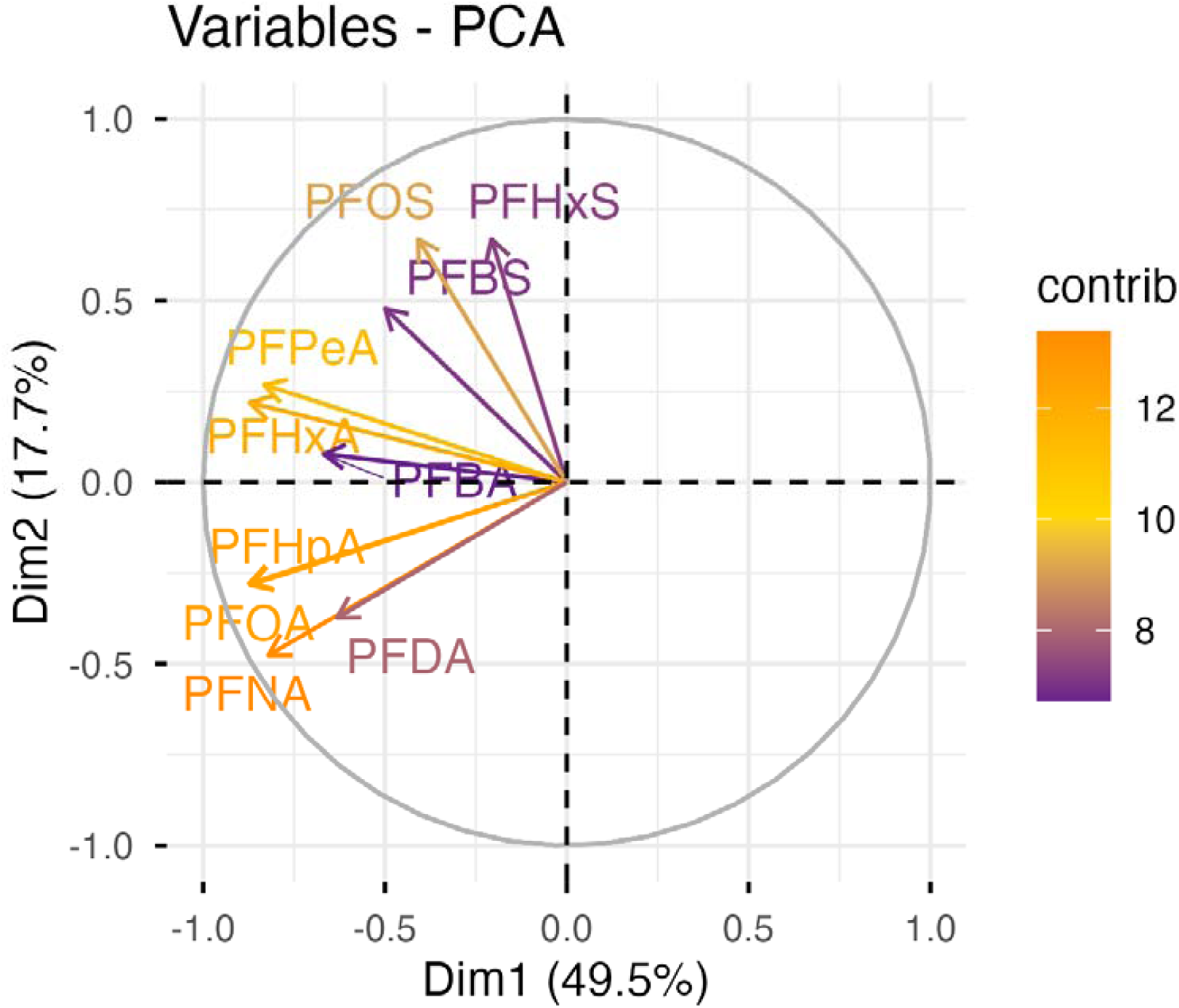
**Principal component analysis of PFAS in water samples from rural wells.**

**Figure S2.**
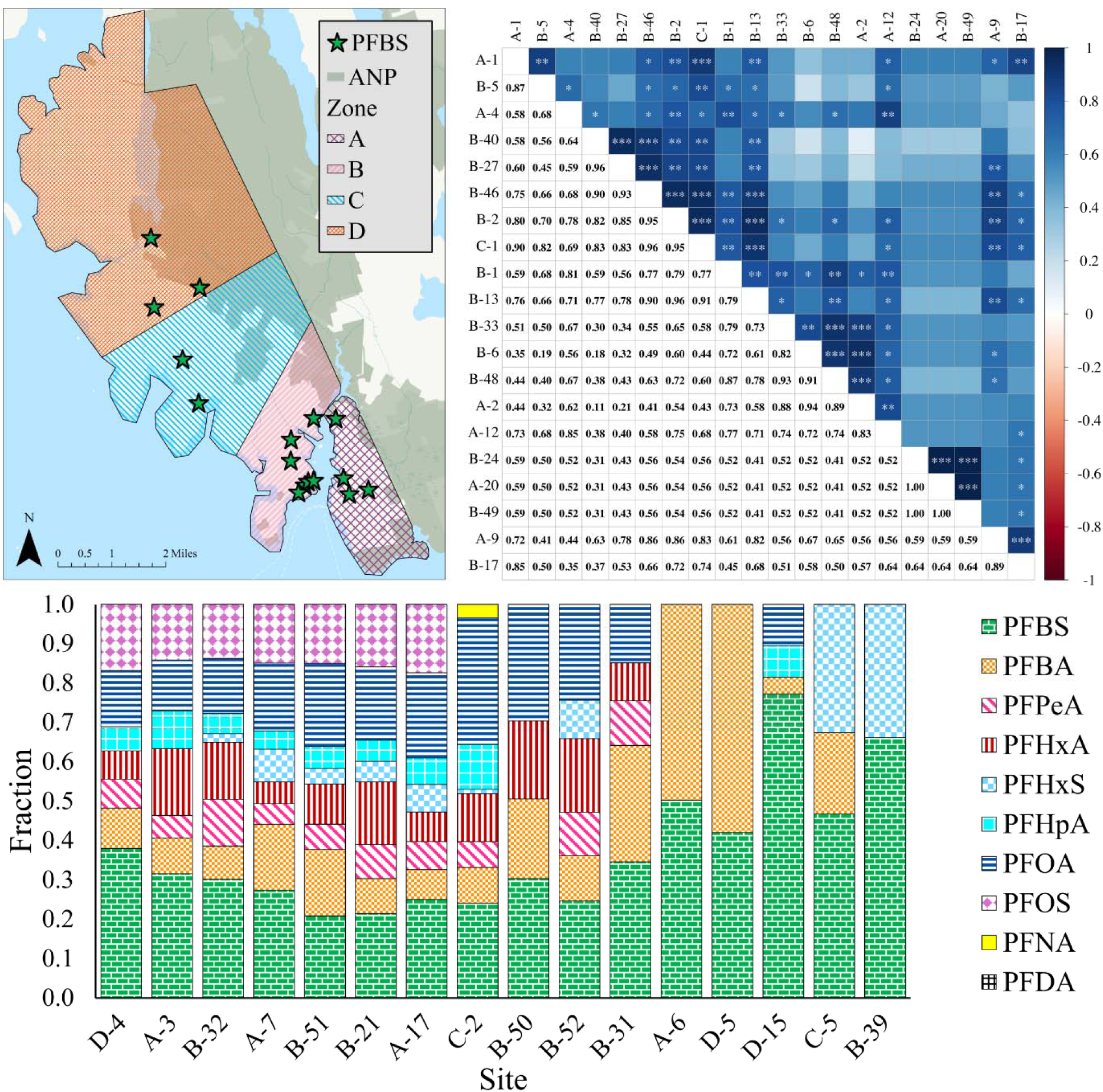
[A] Geographic distribution (as GREEN STARS), [B] pairwise correlations thereof, and [C] fractional chemical profiles of all the samples within the ‘PFBS’ hierarchical cluster illustrated in Figure 2, by the black box surrounding a number of sites in the BOTTOM of the fourth column. ‘ANP’ designates Acadia National Park.

**Figure S3.**
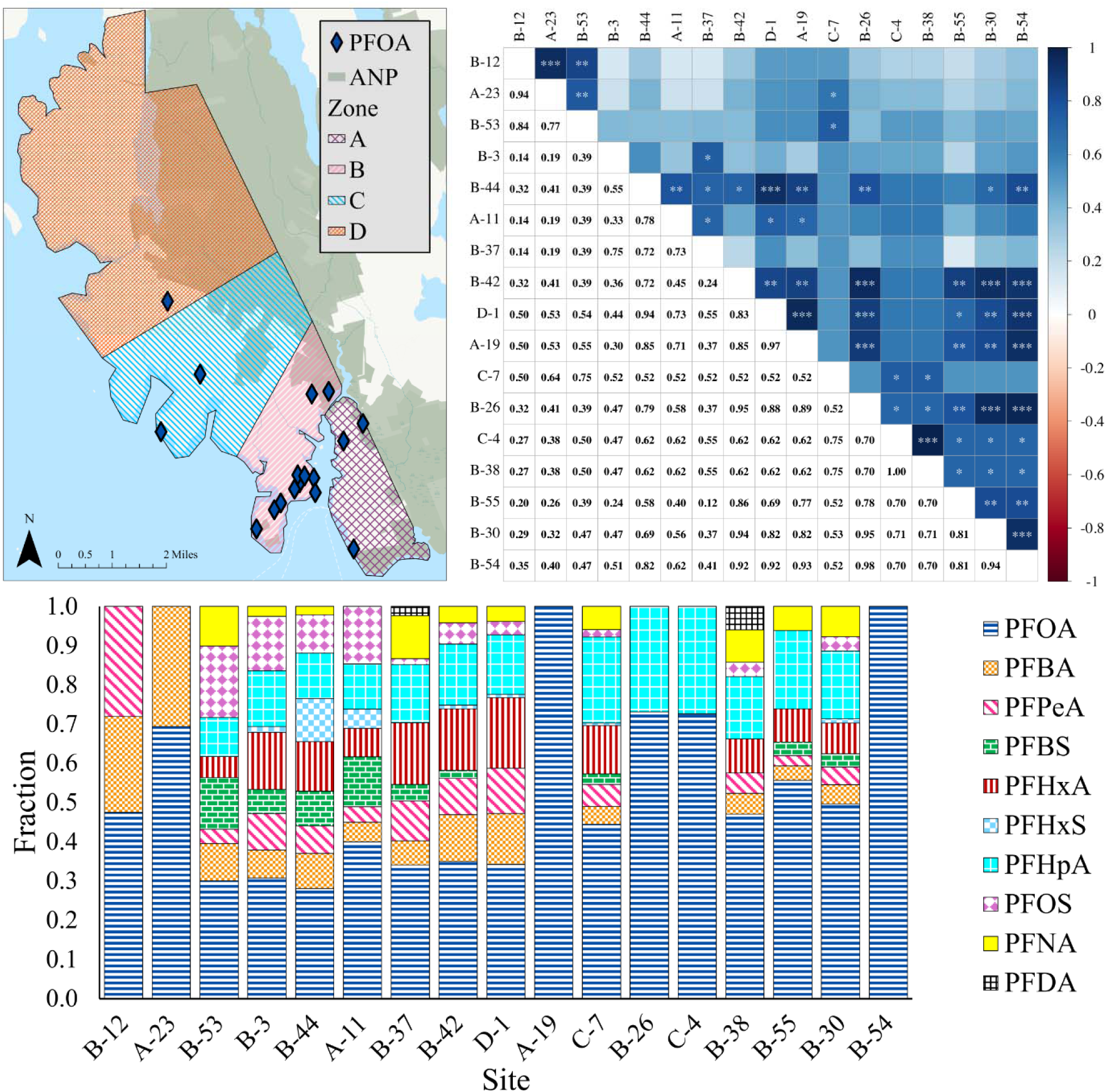
[A] Geographic distribution (as BLUE DIAMONDS), [B] pairwise correlations thereof, and [C] fractional chemical profiles of all the samples within the ‘PFOA’ hierarchical cluster illustrated in Figure 2, by the black box surrounding a number of sites in the BOTTOM of the first column.

**Figure S4.**
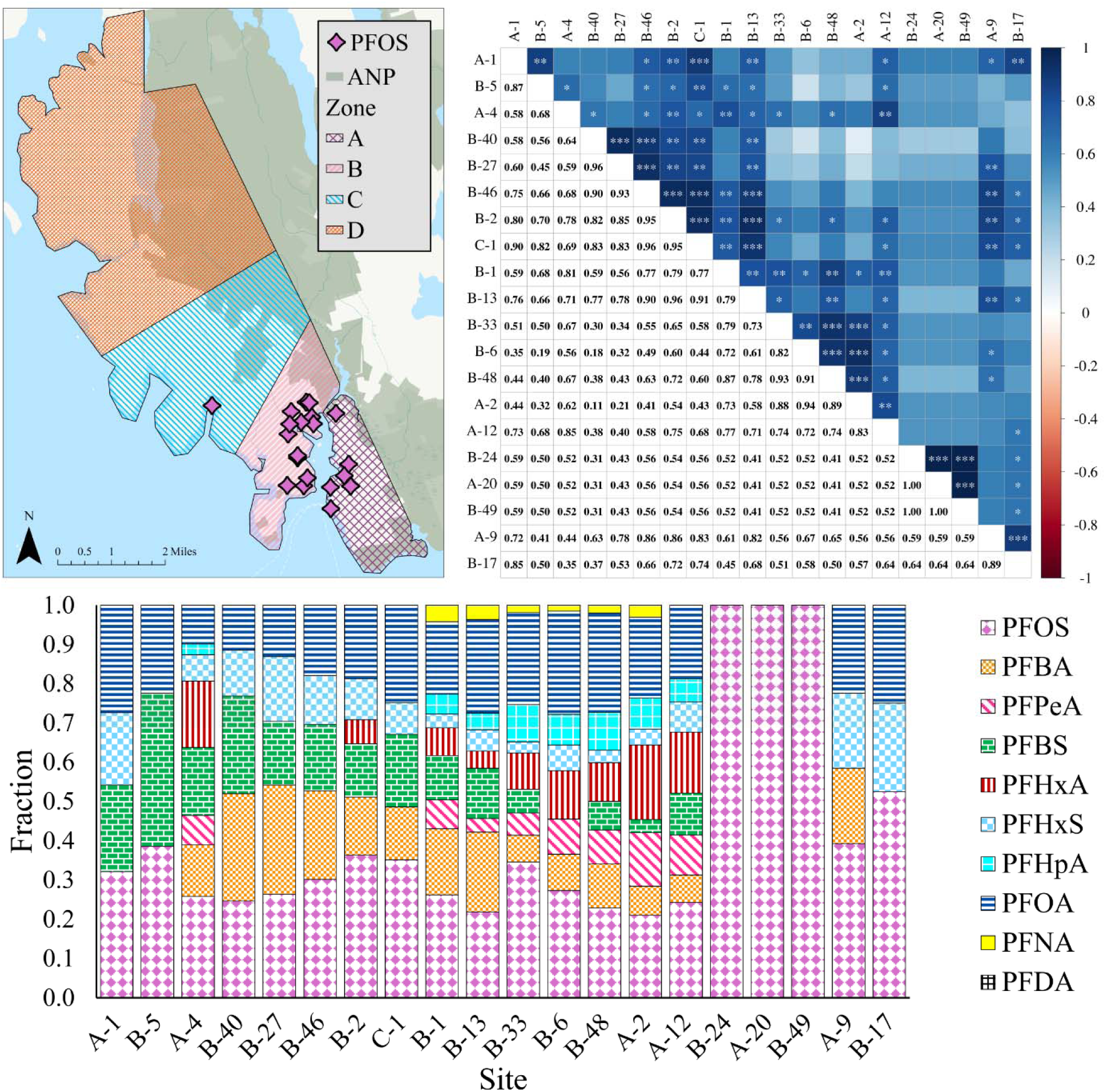
[A] Geographic distribution (as PINK DIAMONDS), [B] pairwise correlations thereof, and [C] fractional chemical profiles of all the samples within the ‘PFOS’ hierarchical cluster illustrated in Figure 2, by the black box surrounding a number of sites in the second column. ‘ANP’ designates Acadia National Park.

**Figure S5.**
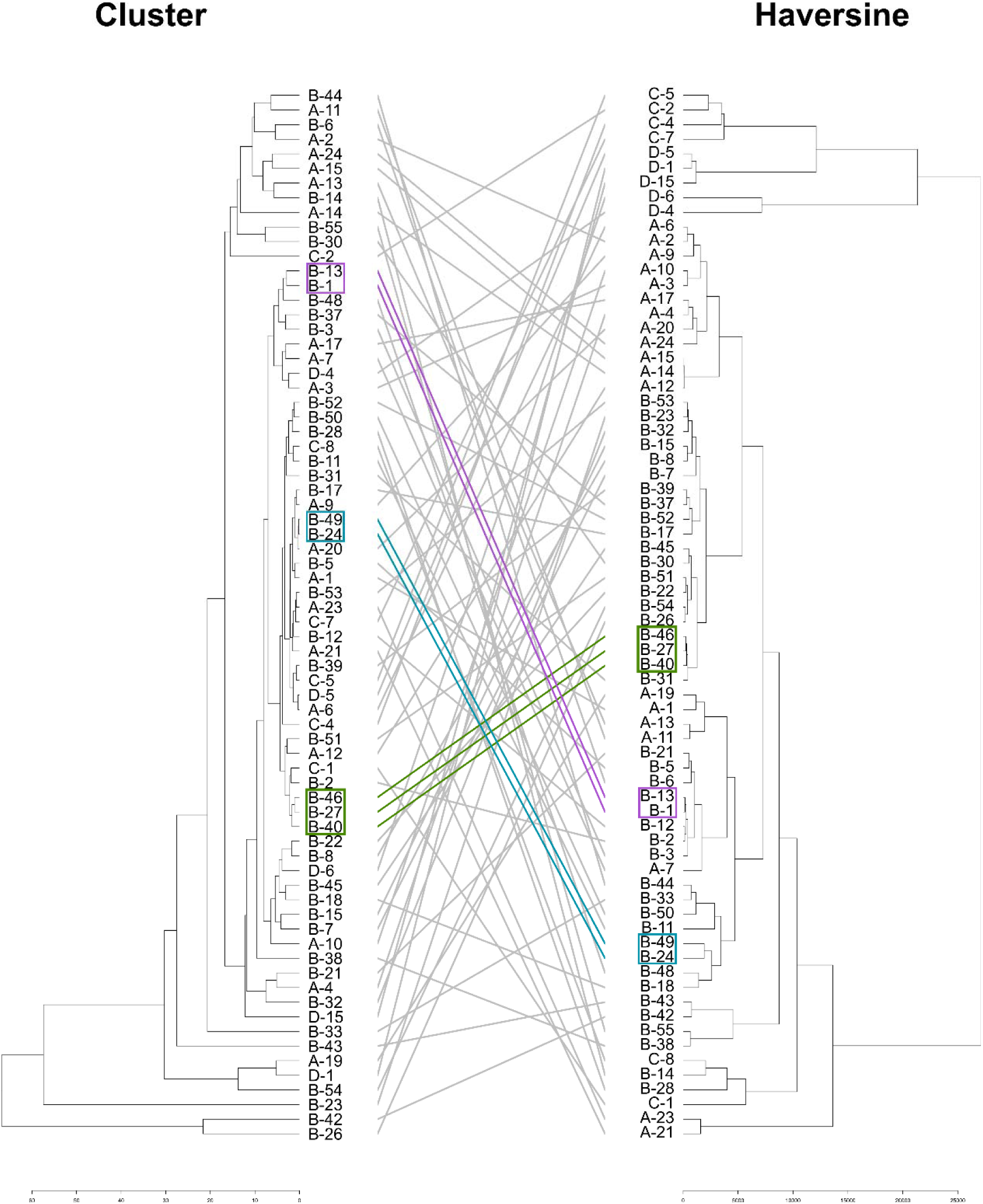
Comparison of hierarchical cluster analyses of the chemical profiles (LEFT) and the Haversine great circle (“as the crow flies”) distance between each site (RIGHT). Colored lines indicate different sets of sites/samples that are adjacent to one another in both chemical and physical space. These all fall in the B Zone. The blue and green lines indicate separate sets of two houses with similar profiles and close proximity. The red lines indicate a set of three houses with similar profiles and close proximity.

**Figure S6.**
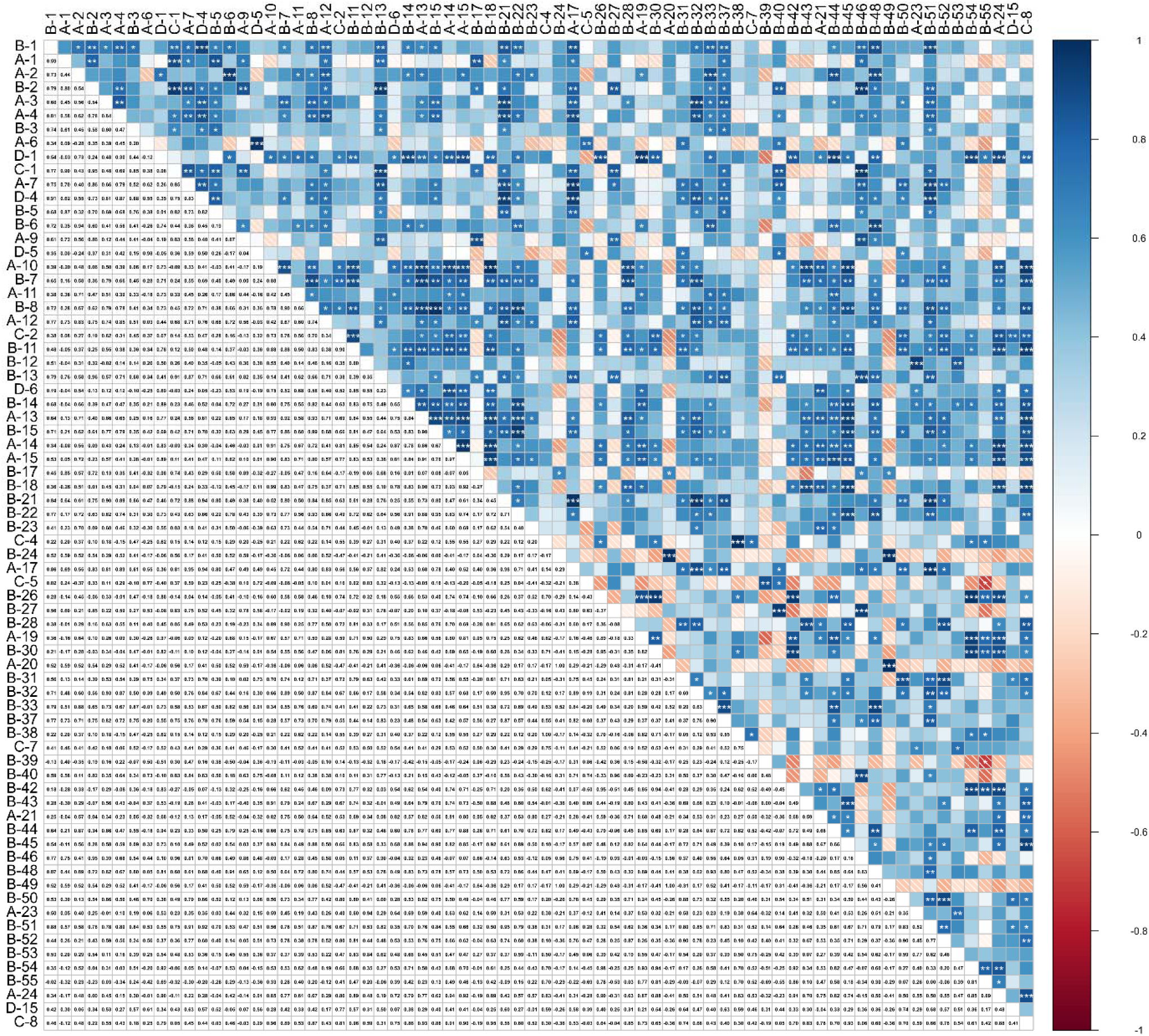
Spearman’s rank correlation for each pair of private wells with detectable PFAS in drinking water. Positive correlations are blue; negative correlations are red. Sites are mirrored across the diagonal (white squares) and ordered along the axes by similarity. *=p **<**0.05, **=p <0.01, ***p <0.001

