## Supplemental Files for "PFAS Profiles in Private Wells Reveal Septic System Sources of Contamination in a Rural Maine Community"

Richard F. Hilliard et al.

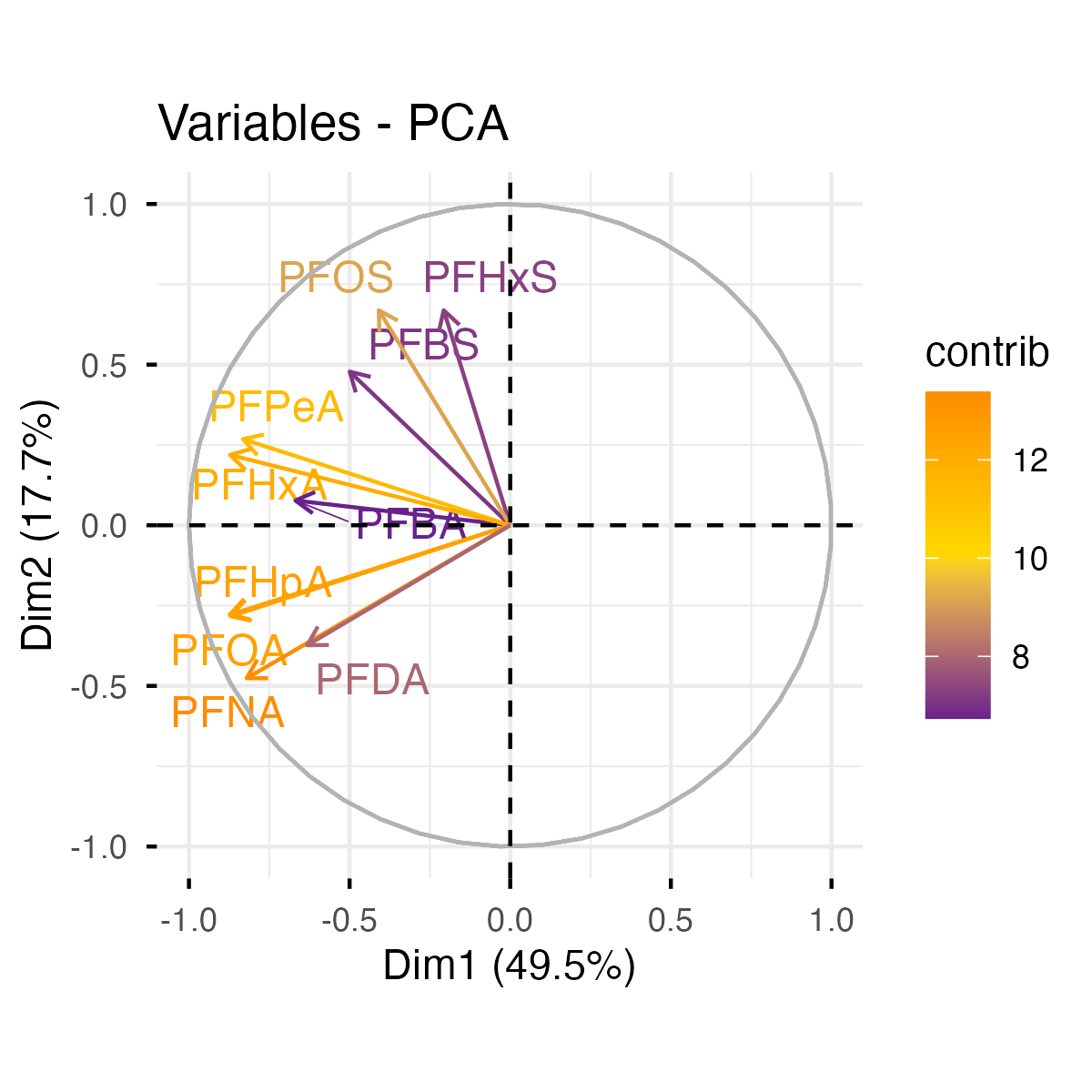

**Figure S1. Principal component analysis of PFAS in water samples from rural wells.**

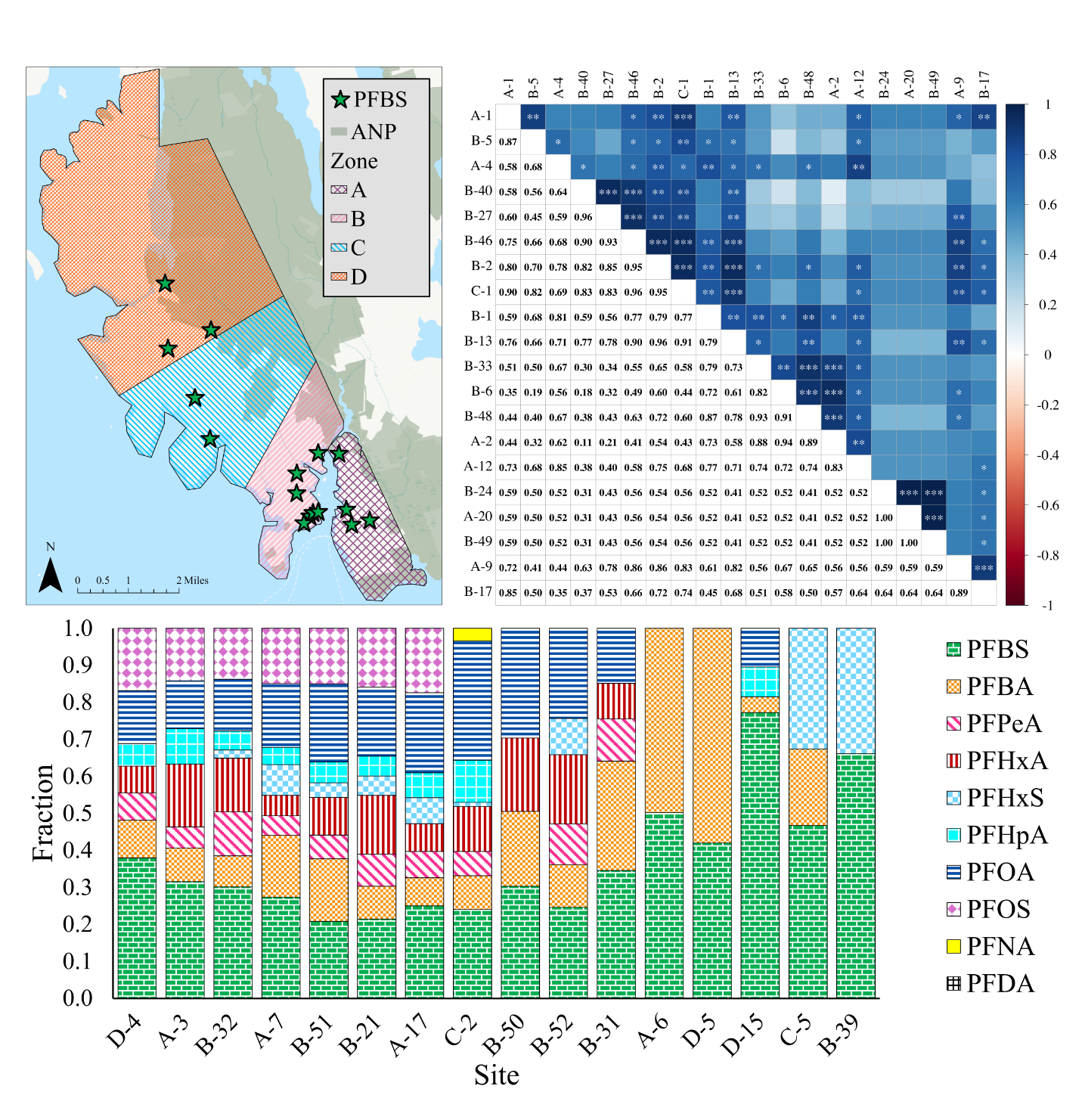

**Figure S2. [A] Geographic distribution (as GREEN STARS), [B] pairwise correlations thereof, and [C] fractional chemical profiles of all the samples within the ‘PFBS’ hierarchical cluster illustrated in Figure 2, by the black box surrounding a number of sites in the BOTTOM of the fourth column. ‘ANP’ designates Acadia National Park.**

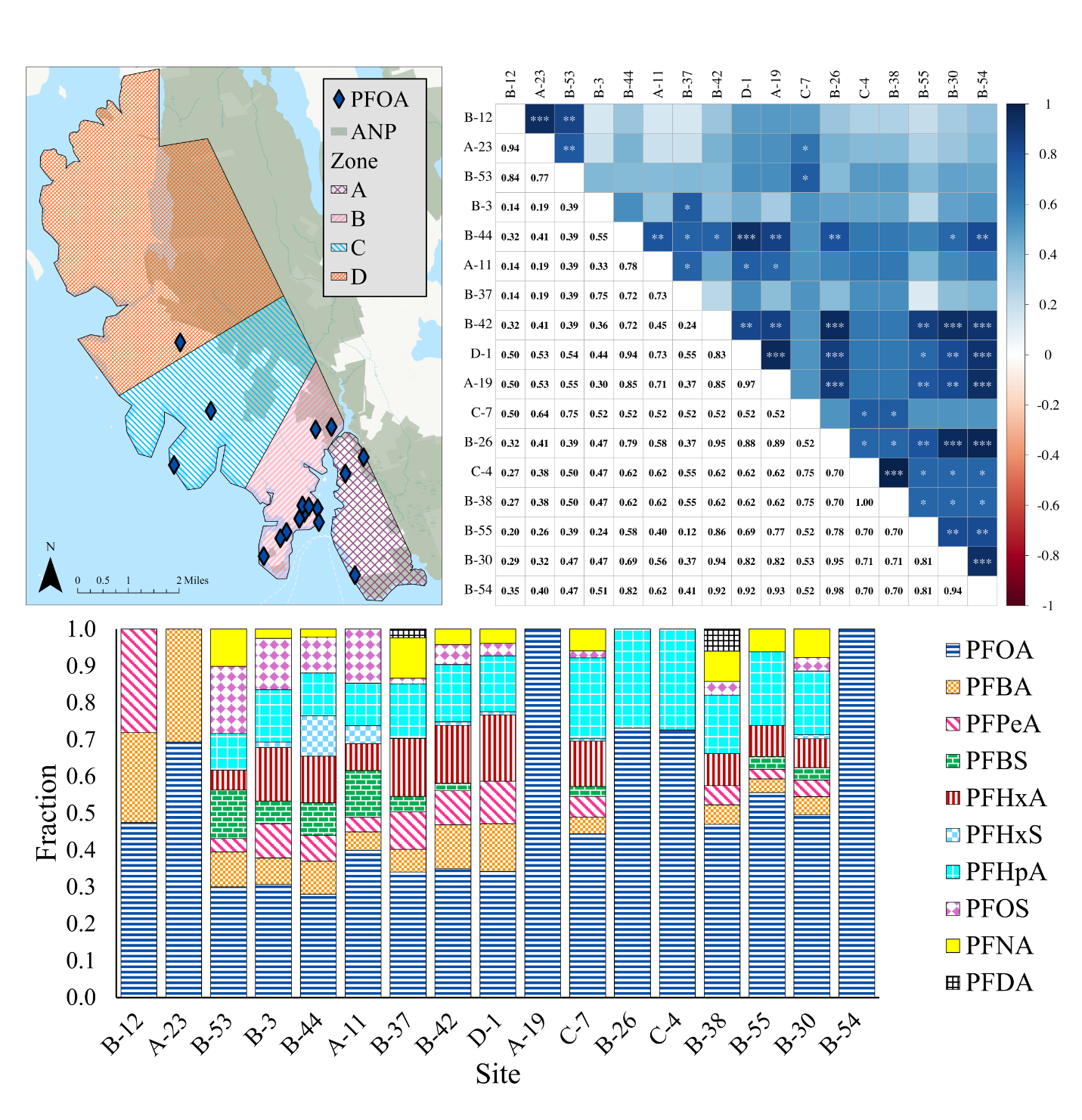

**Figure S3. [A] Geographic distribution (as BLUE DIAMONDS), [B] pairwise correlations thereof, and [C] fractional chemical profiles of all the samples within the ‘PFOA’ hierarchical cluster illustrated in Figure 2, by the black box surrounding a number of sites in the BOTTOM of the first column.**

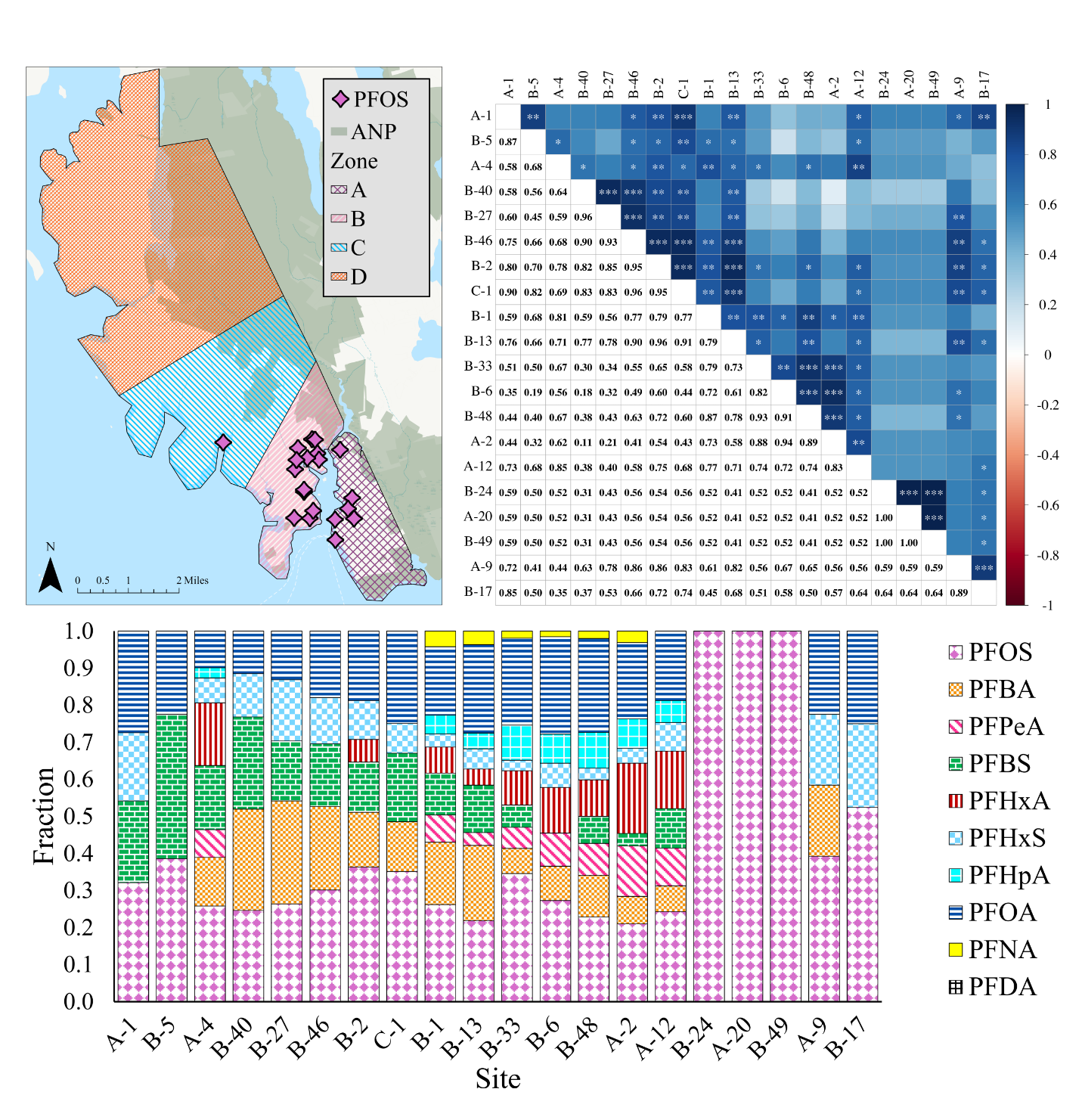

**Figure S4. [A] Geographic distribution (as PINK DIAMONDS), [B] pairwise correlations thereof, and [C] fractional chemical profiles of all the samples within the ‘PFOS’ hierarchical cluster illustrated in Figure 2, by the black box surrounding a number of sites in the second column. ‘ANP’ designates Acadia National Park.**

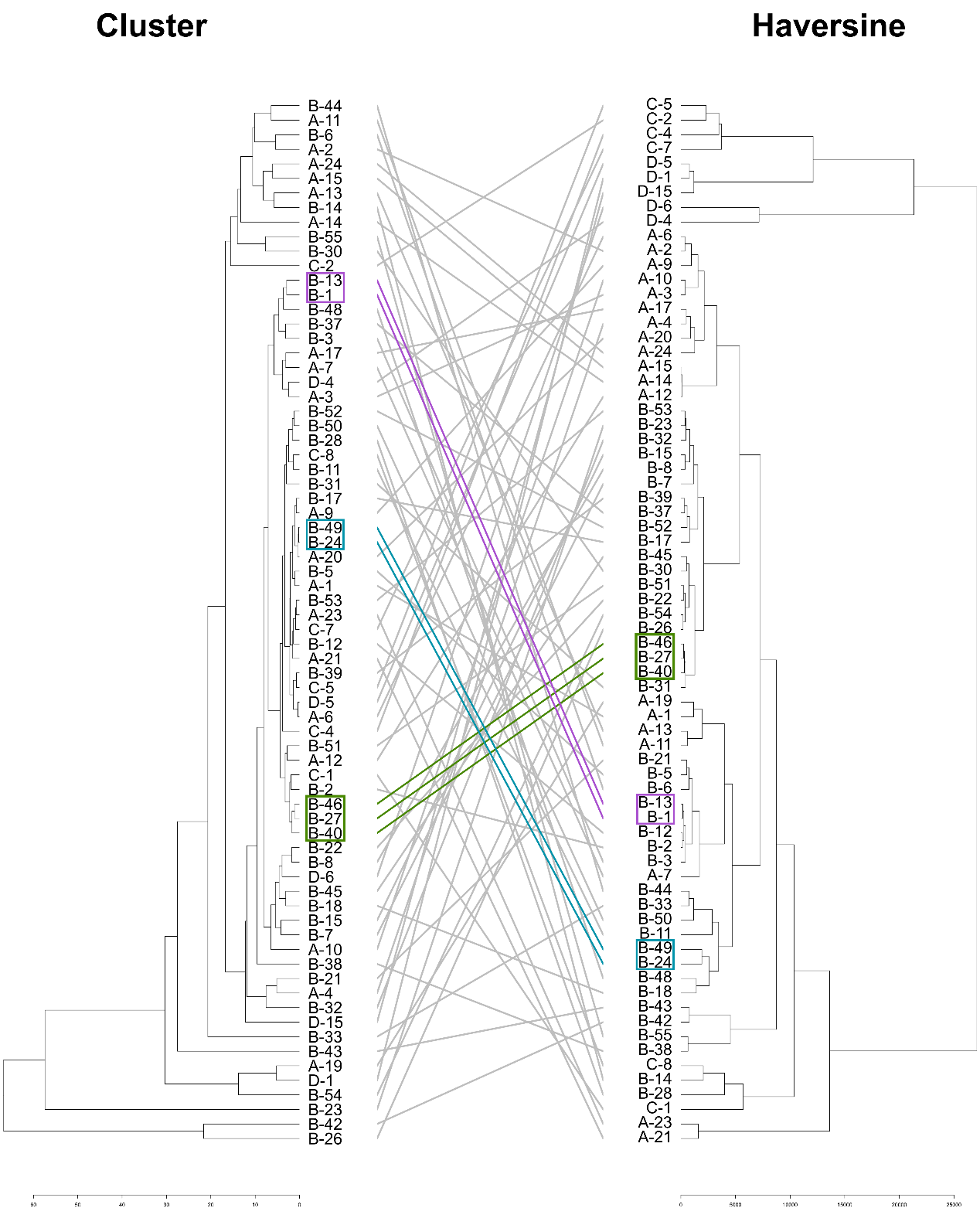

**Figure S5. Comparison of hierarchical cluster analyses of the chemical profiles (LEFT) and the Haversine great circle (“as the crow flies”) distance between each site (RIGHT). Colored lines indicate different sets of sites/samples that are adjacent to one another in both chemical and physical space. These all fall in the B Zone. The blue and green lines indicate separate sets of two houses with similar profiles and close proximity. The red lines indicate a set of three houses with similar profiles and close proximity.**

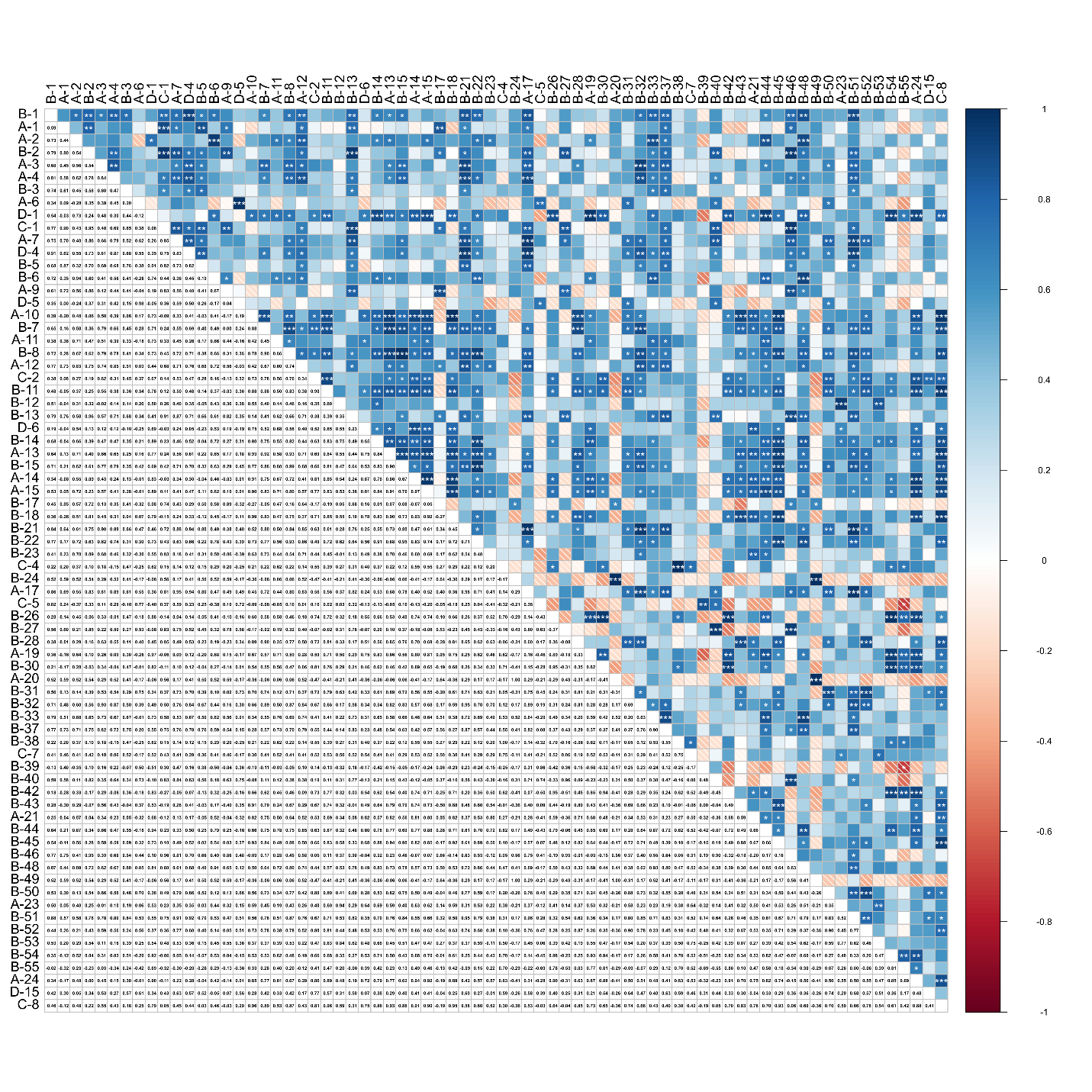

**Figure S6. Spearman’s rank correlation for each pair of private wells with detectable PFAS in drinking water. Positive correlations are blue; negative correlations are red. Sites are mirrored across the diagonal (white squares) and ordered along the axes by similarity. *=p <0.05, **=p <0.01, ***p <0.001**
